# The lost microbes of COVID-19: Bifidobacterium, Faecalibacterium depletion and decreased microbiome diversity associated with SARS-CoV-2 infection severity

**DOI:** 10.1101/2021.09.02.21262832

**Authors:** Sabine Hazan, Neil Stollman, Huseyin Bozkurt, Sonya Dave, Andreas J. Papoutsis, Jordan Daniels, Brad D. Barrows, Eamonn MM Quigley, Thomas J. Borody

## Abstract

**Objective:** The study objective was to compare gut microbiome diversity and composition in SARS-CoV-2 polymerase chain reaction (PCR)-positive patients whose symptoms ranged from asymptomatic to severe, versus PCR-negative exposed controls.

**Design:** Using a cross-sectional design, we performed shotgun next-generation sequencing (NGS) on stool samples to evaluate gut microbiome composition and diversity in both patients with SARS-CoV-2 PCR- confirmed infections, that had presented to Ventura Clinical Trials for care from March 2020 through October 2021, and SARS-CoV-2 PCR-negative exposed controls. Patients were classified as being asymptomatic or having mild, moderate, or severe symptoms based on NIH criteria. Exposed controls were individuals with prolonged or repeated close contact with patients with SARS-CoV-2 infection or their samples, e.g. household members of patients or frontline healthcare workers. Microbiome diversity and composition were compared between patients and exposed controls at all taxonomic levels.

**Results:** Compared with controls (n=20), severely symptomatic SARS-CoV-2 infected patients (n=28) had significantly less bacterial diversity (Shannon Index, P=0.0499; Simpson Index, P=0.0581), and positive patients overall had lower relative abundances of Bifidobacterium (P<0.0001), Faecalibacterium (P=0.0077), and Roseburium (P=0.0327), while having increased Bacteroides (P=0.0075). Interestingly, there was an inverse association between disease severity and abundance of the same bacteria.

**Conclusion:** We hypothesize that low bacterial diversity and depletion of Bifidobacterium genera either before or after infection led to reduced pro-immune function, thereby allowing SARS-CoV-2 infection to become symptomatic. This particular dysbiosis pattern may be a susceptibility marker for symptomatic severity from SARS-CoV-2 infection and may be amenable to pre-, intra-, or post infection intervention.**Keywords:** SARS-CoV-2, COVID, Microbiome, Bifidobacterium, Faecalibacterium, Bacteriodes, Shannon Index, Simpson Index, Severity, Microbiota

**Registration:** clinicaltrials.gov NCT04031469 (PCR -) and 04359836 (PCR+)

|  |  |
| --- | --- |
| **What is already known about this subject?** | The gut microbiome is intrinsically related to host immune response (e.g., inflammation, TH1 vs. TH2) and susceptibility to infection. |
| **What are the new findings?** | Patients with SARS-CoV-2 infections possess significantly less bacterial diversity, lower abundance of *Bifidobacterium* and *Faecalibacterium*, and increased abundance of *Bacteroides* at the genus level compared with SARS-CoV-2 exposed controls. There are inverse associations between disease severity and the Shannon and Simpson diversity indices and also with *Bifidobacterium* and *Faecalibacterium* abundance. There is also a direct association between severity and *Bacteroides* abundance. |
| **How might it impact on clinical practice in the foreseeable future?** | Boosting of *Bifidobacterium* or *Faecalibacterium* through probiotic supplementation or fecal microbiota transplant is worthy of exploration in the management of patients with acute severe disease or protracted infection. If the changes that we document precede SARS-CoV-2 infection in those who are most severely affected, this therapeutic approach may be of particular interest. Conversely, if the reduction follows infection, then repopulation of the gut microbiome may reduce long-term effects related to gut microbiome composition changes with SARS-CoV-2 infection. |

## INTRODUCTION

The abundance of *Bifidobacterium* decreases with increasing age and BMI[1] and *Bifidobacterium* is the active ingredient of many probiotics. *In-vitro* studies have demonstrated the benefits of these Gram positive bacteria to include enhanced ATP production, immune modulation and competence,[2, 3, 4, 5, 6, 7, 8] mucosal barrier integrity, restriction of bacterial adherence to, and invasion of, the intestinal epithelium, and modulation of central nervous system activity.[9, 10] Additionally, many *Bifidobacterium spp.* have anti-inflammatory properties: *Bifidobacterium animalis, Bifidobacterium longum,* and *Bifidobacterium bifidum* decrease the function of the “master switch[2]” pro-inflammatory tumor necrosis factor- (TNF-) α, increase the anti-inflammatory cytokine interleukin- (IL-10), and promote the TH1 while inhibiting the TH2 immune response.[8] In a mouse model of inflammatory bowel disease, *B. bifidum* and *B. animalis* reduced pro-inflammatory cytokines and restored intestinal barrier integrity.[8] With respect to SARS-CoV-2 infection, there is immunologic coordination between the gut and lungs.[11, 12, 13] Numerous studies have suggested that a healthy gut microbiome may be associated with a decrease in SARS-CoV-2 related mortality[14] and that probiotics should be considered for prophylaxis[15] and/or treatment of SARS-CoV-2 or its associated secondary infections. [15][16] [17] However, as of Feb. 2022, despite the publication of nearly 8000 studies on SARS-CoV-2 infection, few ongoing studies (clinicaltrials.gov: NCT04443075 and NCT04486482) and only 5 publications to date have examined gut microbiome changes in SARS-CoV-2 infected patients. Nevertheless, an association between the status of the gut microbiome and outcome from this infection has been suggested. Accordingly, increased abundance of the *Streptococcus, Rothia, Veilonella,* and *Actinomyces* genera was associated with inflammation, [18] whereas increased abundances of *Collinsella aerofaciens, Collinsella tanakaei, Streptococcus infantis*, and *Morganella morganii* were associated with fecal samples with high SARS-CoV-2 infectivity,[19] and increased Lachnospriaceae and Enterobacterioaceae abundances were associated with increased mortality and need for artificial ventilation.[19] Species potentially protective against SARS-CoV-2 infection include *Parabacteroides merdae, Bacteroides stercoris, Alistipes onderdonkii, Lachnospiracea bacterium*[19] and *F. prausnitzii.*[19, 20], while vulnerability to infection and increased severity were associated with decreased abundance of *B. bifidum*.[20, 21] A recent study correlated aspects of the gut microbiome with “Long-COVID” including reduced levels of F. prausnitzii on admission[22]. In short, there is still a compelling need to elucidate changes in the human gut microbiome due to SARS-CoV-2 and their relationship with clinical outcomes.

The scientific community and lay public are increasingly interested in the therapeutic potential of probiotics. *Bifidobacteria* spp. have potential to improve clinical conditions ranging from inflammatory bowel disease (IBD)[23] to *Clostridioidies difficile* infections.[23, 24, 25, 26] Treatment with specific strains of *Bifidobacteria in vitro* have been shown to reduce toxins from *C. difficile .*[25] *In vivo*, *Bifidobacteria* can restore colonic integrity,[27] and *B. longum* administered intranasally in mice prior to exposure to influenza has been associated with reduction in mortality.[4] Given that *Bifidobacteria* are a common component of several probiotic products and appear to be associated with SAR-CoV-2 infections, one could ask if probiotics might have a role in SARS-CoV-2 therapy or prevention.

Herein, we evaluate the relationships between gut microbiome diversity and composition compared to clinical outcome in cross-sectional groups of SARS-CoV-2 PCR-confirmed positive patients (ranging from asymptomatic to severely symptomatic) versus SARS-CoV-2 PCR-confirmed negative controls. Our controls are SARS-CoV-2 exposed persons who remained PCR-negative and asymptomatic. The controls likely had similar viral exposures, but appeared protected against infection, and our data suggests some protection may reside in the microbiome.

## METHODS

### Study Design and Patients

Individuals who were tested for SARS-CoV-2 infection either because they were symptomatic or had been exposed to a “case” were eligible for enrollment the week following testing if either they or a household member was positive. Controls eligible for enrollment were PCR-negative for SARS-CoV-2, remained antibody negative for 3 to 6 months, and asymptomatic for 6 to 12 months. Additionally, controls had to either share a household with at least one symptomatic SARS-CoV-2-positive individual or be a healthcare worker who had been repeatedly exposed to symptomatic SARS-CoV-2-positive patients or numerous SARS-CoV-2 positive samples. All exposed controls were ones that, despite exposure to SARS-CoV-2, chose not to quarantine or take prophylaxis for SARS-CoV-2 infection and none had yet been vaccinated.

Patients did not wear PPE inside their homes and staff did not wear *full* PPE (i.e., did not wear masks) at the office because of its scarcity during this global pandemic. Patients undergoing treatment with total parenteral nutrition, or those with a history of significant gastrointestinal surgery (e.g. bariatric surgery, total colectomy with ileorectal anastomosis, proctocolectomy, postoperative stoma, ostomy, or ileoanal pouch) were excluded.

This study was performed between March 1st, 2020 to October 31^st^, 2021, with all but one subject recruited prior to June 1^st^, 2021. During that time, alpha and epsilon variants predominated in the United States[28].

The study was conducted in accordance with ethical principles of the Declaration of Helsinki, the International Council for Harmonisation (ICH) Harmonised Tripartite Guideline for Good Clinical Practice (GCP), and the Ethical and Independent Review Board (IRB). All patients provided written informed consent to participate.

### Assessments

A self-administered questionnaire solicited information on symptom severity, past medical history, current medication and probiotic use, and exposure to recreational drugs or animals. Patients with SARS-CoV-2 infection were further classified as being either asymptomatic carriers or having mild, moderate, or severe symptoms as per National Institute of Health, Clinical Spectrum of SARS-CoV-2 Infection criteria.[29, 30] Asymptomatic PCR-confirmed SARS-CoV-2 positive household members of SARS-CoV-2 infected patients were categorized as asymptomatic carriers. Patients and controls were classified as underweight, normal weight, overweight, obese, or severely obese based on body mass index criteria of the Center for Disease Control and Prevention.[31]

### Stool Sample Collection and Processing

Patients and controls within the same household collected stool samples within a week of the index case being positive. Patients had stool samples collected at baseline, prior to any treatment, and within 48 hrs. of symptom onset. No subjects had been commenced on antibiotics, SARS-CoV-2 infection treatments, OTC remedies (e.g. vitamins, antipyretics, analgesics), or supplemental oxygen between the time of symptom onset or demonstration of PCR positivity and stool collections. Subjects were instructed (and educated on the procedure and sterile methods) to collect 1 mL of fresh stool and place it directly in a DNA/RNA Shield^TM^ Fecal Collection Tube (Zymo Research, Tustin, CA, USA), and then mix sample thoroughly. One ml of feces is more than sufficient to capture the microflora of the gut accurately and consistently. This method is chosen to eliminate the need for whole stool mixing and aliquoting. The solution in the Fecal Collection Tube is designed to preserve samples at ambient temperature (4°C-25°C) for > 2 years, or below - 20°C indefinitely. Once samples reached our laboratory, they were immediately frozen at -20°C.

Following fecal collection, each individual sample DNA was extracted and purified with the Qiagen PowerFecal Pro DNA extraction kit. The isolated DNA was then quantitated utilizing the Quantus Fluorometer with the QuantFluor ONE dsDNA kit. After DNA quantification, the DNA was normalized, i.e. all samples begin library preparation (following DNA extraction and purification) with 100 ng input of DNA. Libraries were then prepared using shotgun methodology with Illumina‟s Nextera Flex kit. Samples then underwent the shotgun metagenomic processing procedure of tagmentation, amplification, indexing, and purification. Following completion of this shotgun metagenomic standard protocol, purified libraries were again normalized to standardize sequencing depth during the NGS run on the NextSeq 500/550. We achieved consistency of sequencing depth (ie number of reads) by normalizing the samples‟ pooling concentrations (ie molarity), loading the same number of samples per sequencing run, consistently using the same NextSeq High Output kits.

After completion of sequencing on the Illumina NextSeq with 500/550 High-Output Kits v2.5 (300 cycles), the raw data was streamed in real-time to Illumina‟s BaseSpace cloud for FASTQ conversion. The FASTQ files were then sent through One Codex‟s bioinformatics pipeline for metagenomic annotation and analyses to elucidate the microflora composition and relative abundances of the top genera and species for all patients and controls.

### Data Analysis

We assessed differences in relative abundance across taxa between the gut microbiome of SARS-CoV-2 infected patients and exposed controls and calculated Shannon and Simpson alpha diversity indices with One Codex‟s bioinformatics analysis pipeline utilizing Jupyter Notebook in Python. Specifically, the One Codex Database consists of ∼114K complete microbial genomes (One Codex, San Francisco, CA, USA). During processing, reads were first screened against the human genome, then mapped to the microbial reference database using a k-mer based classification. Individual sequences (NGS read or contig) were compared against the One Codex Database (One Codex) by exact alignment using k-mers, where k = 31(43, 44). Based on the relative frequency, unique k-mers were filtered to control for sequencing or reference genome artifacts.

The sequencing depth followed ProgenaBiome‟s standard operating procedures and was 8,239,475 reads on average for this study. One should note that shallow metagenomic sequencing is typically only 0.5 million reads but is still considered sufficient for taxonomic phyla level analysis (and even genera for the most abundant bacteria).

The relative abundance of each microbial taxonomic classification was estimated based on the depth and coverage of sequencing across every available reference genome. Beta-diversity was calculated as weighted UniFrac distance visualized in a distance matrix using the phylum-level relative abundance obtained from One Codex. 13 genera were selected based on our experience and knowledge of critical players in the gut microbiome, as well as similarity to other studies [18, 20][29] To compare patients across subgroups and patients to exposed controls, ANOVA, Mann-Whitney U, Kruskal-Wallis tests, and chi-square test statistics were conducted using GraphPad version 8 with P-values < 0.05 considered as significant. Dunn‟s *post-hoc* was used for Kruskal-Wallis test, with correction for multiple comparisons in all situations.

All authors had access to study data and reviewed and approved the final manuscript.

## RESULTS

### Patient Characteristics

Demographic and clinical characteristics of patients (n=50) and exposed controls (n=20) are presented in **Supplementary Table S1** and **Supplementary Table S2,** and summarized in **Table 1**. All patients were resident of USA, with states indicated in Supplementary Table 1. 24 of 50 (48%) of patients and 7 of 20 (35%) of exposed controls were male. The mean ± SEM age in years was 50.0 ± 2.5 for patients and 44.4 ± 3.6 for exposed controls. 44 of 50 (88%) of patients were Non-Hispanic White; 5 of 50 (10%), Hispanic; and 1 of 50 (2%), Native American and 17 of 20 (85.0%) of exposed controls were non-hispanic white; 2 of 20 (10.0%), Hispanic; and 1 of 20 (5.0%), Black. Of patients, 28 of 50 (56%) had severe, 12 of 50 (24%) had moderate and 6 of 50 (12%) had mild disease, and 4 of 50 (8%) were asymptomatic. 32 of 50 (64%) of patients and 12 of 20 (60.0%) of exposed controls had underlying comorbidities considered risk factors for increased severity of SARS-CoV-2 infection by the CDC.[1] The mean ± SEM BMI of the 46 patients with available data was 27.1 ± 0.98 compared with 25.1 ± 0.96 for the 20 exposed controls. There was no significant difference (P>0.2) in gender, age, racial demographics, loss of appetite, change in stool frequency, diet, or presence of underlying comorbidities.

**Table 1:**
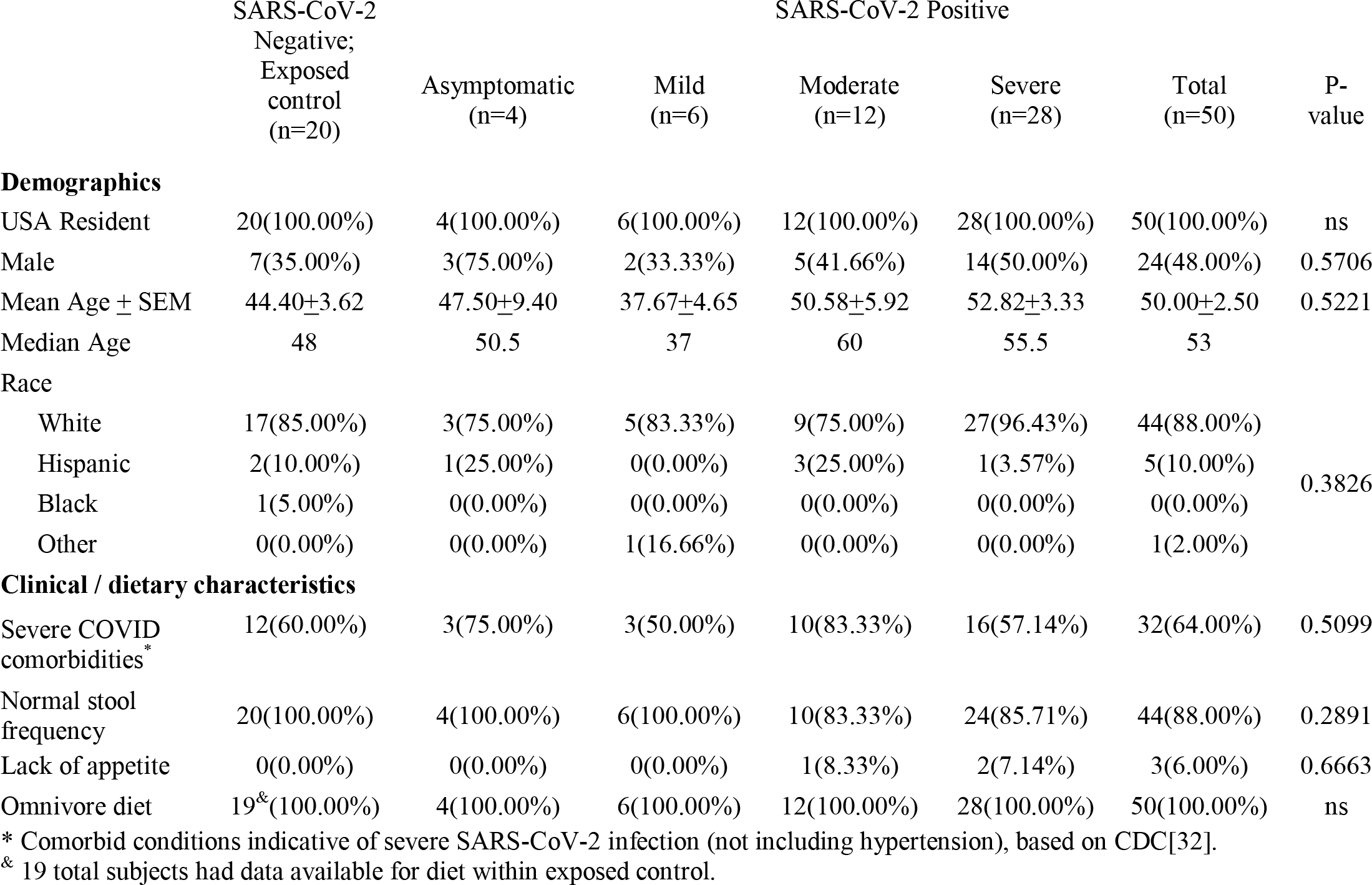
Summary of demographics along with clinical and dietary characteristics of subjects. Numbers in cells indicate number of subjects with percentage in categories, except for age and BMI which indicate value. P-values calculated via one-way ANOVA (age and BMI) or chi-square (others). Normal stool frequency is defined as absence of diarrhea. Total refers to sum of SARS-CoV-2 positive subjects, and it is not used in statistics.

**Table 2:**
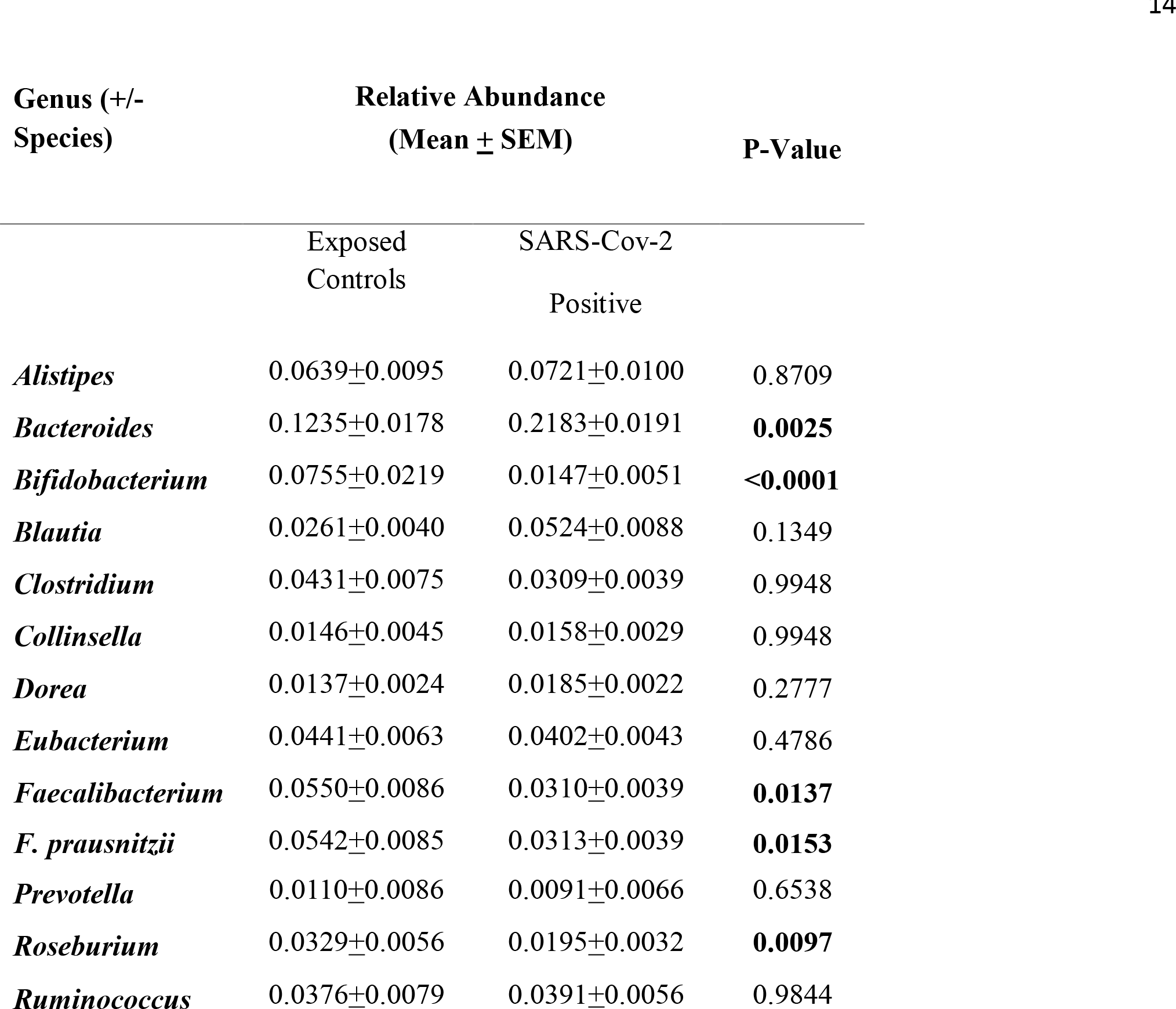
Relative abundances of Bacteroides increase and of Bifidobacterium, Faecalibacterium, and Roseburium decrease in SARS-CoV-2 positive subjects vs. SARS-CoV- 2 negative exposed controls. Mean ± SEM Relative abundances, as well as P-Value via Mann- Whitney-U test are indicated, with bold marking P<0.05.

| Genus (+/-<br>Species) | Relative Abundance<br>(Mean ± SEM) | | P-Value |
| --- | --- | --- | --- |
|  | Exposed<br>Controls | SARS-Cov-2<br>Positive |  |
| *Alistipes* | 0.0639±0.0095 | 0.0721±0.0100 | 0.8709 |
| *Bacteroides* | 0.1235±0.0178 | 0.2183±0.0191 | **0.0025** |
| *Bifidobacterium* | 0.0755±0.0219 | 0.0147±0.0051 | **<0.0001** |
| *Blautia* | 0.0261±0.0040 | 0.0524±0.0088 | 0.1349 |
| *Clostridium* | 0.0431±0.0075 | 0.0309±0.0039 | 0.9948 |
| *Collinsella* | 0.0146±0.0045 | 0.0158±0.0029 | 0.9948 |
| *Dorea* | 0.0137±0.0024 | 0.0185±0.0022 | 0.2777 |
| *Eubacterium* | 0.0441±0.0063 | 0.0402±0.0043 | 0.4786 |
| *Faecalibacterium* | 0.0550±0.0086 | 0.0310±0.0039 | **0.0137** |
| *F. prausnitzii* | 0.0542±0.0085 | 0.0313±0.0039 | **0.0153** |
| *Prevotella* | 0.0110±0.0086 | 0.0091±0.0066 | 0.6538 |
| *Roseburium* | 0.0329±0.0056 | 0.0195±0.0032 | **0.0097** |
| *Ruminococcus* | 0.0376±0.0079 | 0.0391±0.0056 | 0.9844 |

Of the exposed controls, 16 were household contacts of SARS-CoV-2 positive patients in the study, 2 were healthcare workers with extensive, non-protected, exposure to SARS-CoV-2 positive patients, and 2 were laboratory personnel exposed to thousands of SARS-CoV-2 samples (healthcare workers and laboratory personnel did not wear full PPE, i.e., did not wear a face mask, due its scarcity; see methods). During the timeframe of the study, none of the patients or controls were on SARS-CoV-2 prophylaxis or treatment, and none had yet been vaccinated. No patients or exposed control were positive for SARS-CoV-2 prior to the study.

### Gut Microbiome Diversity and Composition

**Figure 1** depicts pie charts of the composition of the gut microbiome for the exposed control at the phylum level (**Figure 1A**) and genus level (**Figure 1B**). At phylum level, Firmicutes and Bacteroides dominated, comprising 59.6% (exposed control) and 54.7% (SARS-Cov-2 positive) and 29.1% (Exposed control) and 40.4% (SARS-Cov-2 positive) of all phyla, repectively. At the level of genus, *Bacteroides* contributed 12.4% (exposed control) and 21.8% (SARS-Cov-2 positive), *Alistipes* 6.4% (Exposed control) and 7.2% (SARS-Cov-2 positive), and *Bifidobacteria* 7.6% (exposed control) and 1.5% (SARS-Cov-2 positive).

**Figure 1:**
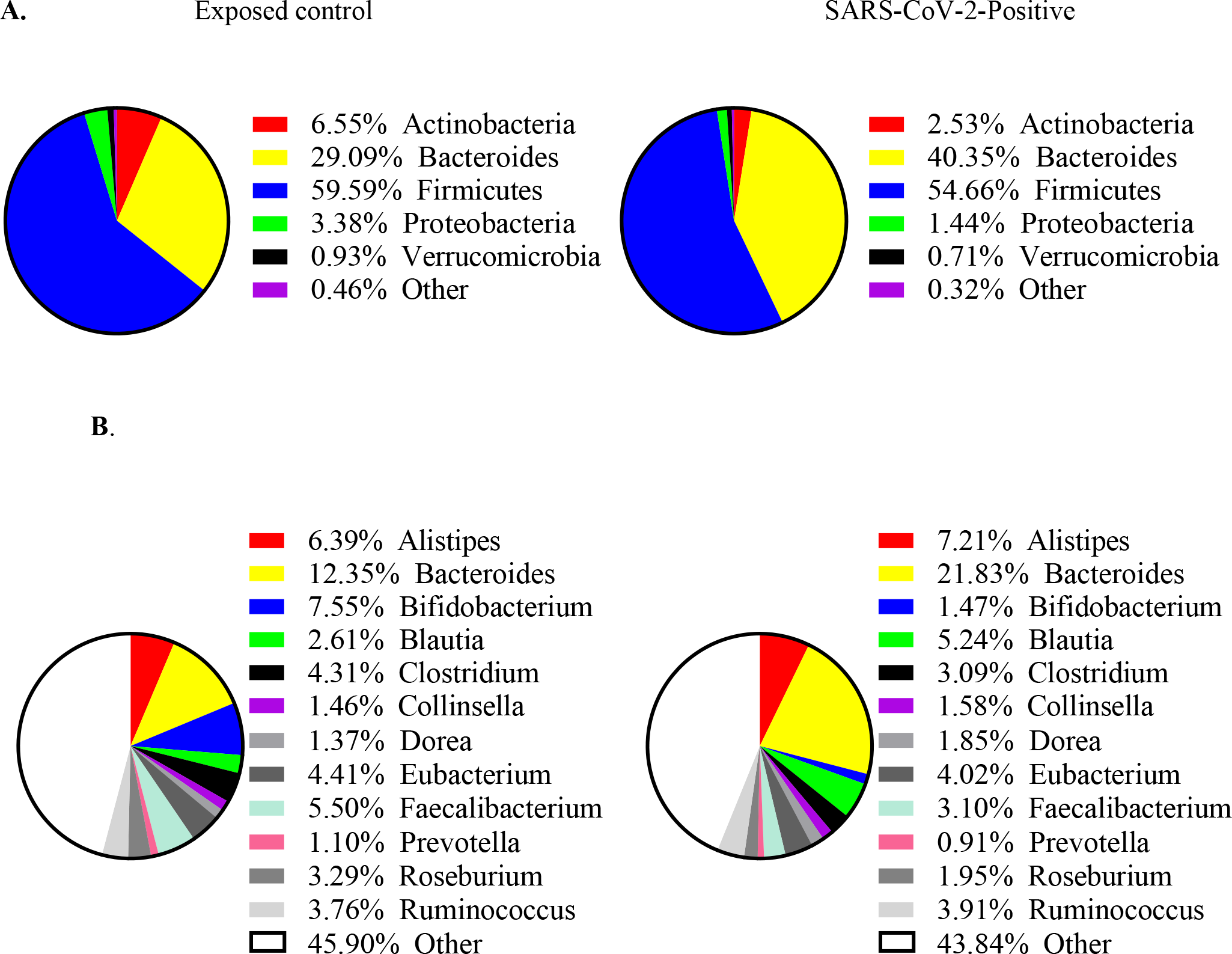
Distribution of bacterial relative abundance in various A. phlya and B. genera for exposed control (n=20, left) and SARS-CoV-2 positive subjects (n=50, right).

**Figure 2** shows two diversity indices for all subgroups studied, namely Shannon Diversity (**Figure 2A**) and Simpson Diversity index (**Figure 2B**). The overall P value for Shannon index (richness of bacterial composition) demonstrated a significant (P=0.0499) decrease in diversity with increased severity, and significance was seen for exposed ctrl vs. severely symptomatic (P=0.0201), analyzed via. Kruskal-Wallis test. The Simpson (evenness of bacterial composition) indexes showed a trend (P=0.0581) of a decrease in diversity with increased SARS-CoV-2 severity.

**Figure 2:**
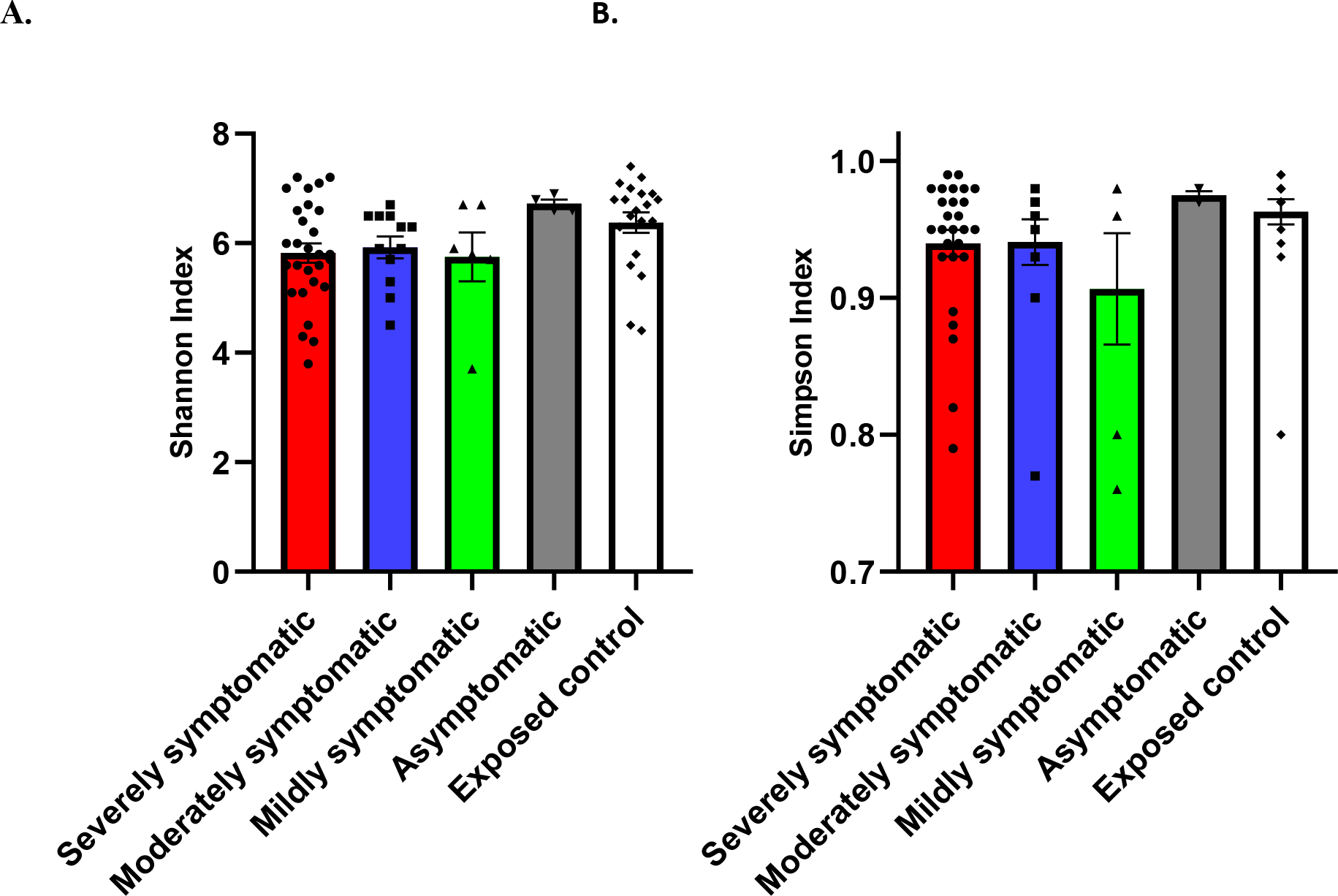
Diversity of gut microbiome composition of SARS-CoV-2 positive patients. (Severely symptomatic: n=28; Moderately Symptomatic: n=12; Mildly Symptomatic: n = 6; Asymptomatic: n=4) vs. exposed controls (n=20). A. Shannon index (P=0.0499) B. Simpson index (P=0.0581). Differences between severely symptomatic positive and exposed negative controls were analyzed via. Kruskal-Wallis test Dunn‟s post-hoc, correcting for multiple comparisons, showing significant for Shannon index at P=0.0201.

Further metagenomic analysis comparing SARS-CoV-2 patients and controls revealed significant differences in relative abundance of specific bacteria. The relative abundance of SARS-COv-2 positive (exposed control) vs. negative subjects is presented in **Table 2**, along with comparative P- Values via. Mann-Whitney-U test. Patients with SARS-CoV-2 infection showed a significantly decreased relative abundance of *Bifidobacterium* and *Faecalibacterium*, and significantly increased relative abundance of *Bacteroides* (**Table 2**)

**Table 3A** lists the genera / species relative abundances (mean±SEM) for various levels of severity of SARS-CoV-2 positive patients vs. exposed control. Analyzed via Kruskal-Wallis test, the main effect (i.e. overall P-value) of these changes are shown in the left column. **Table 3B** proceeds to compare, correcting for multiple comparison, the three levels of severity in infected patients vs. exposed control and asymptomatic groups. Specifically, increased disease severity was associated with decreased relative abundance of *Bifidobacterium, Faecalibacterium, F. prausnitizii,* and *Roseburium,* along with an increased relative abundance of *Bacteroides*.

**Table 3:** Relative abundance of Bacteroides increases and that of Bifidobacterium, Faecalibacterium and Roseburium decreases with increasing severity of disease. A. Relative abundance of genera/species for various severity of SARS-Cov-2 positivity, as well as exposed controls (SARS-CoV-2 negative). Overall P-value of Kruskal-Wallis test is indicated in „Main Effect‟ column. B. P-value for various levels of severity of symptomatic infection, compared to negative Exposed Controls and positive asymptomatic subjects, using Dunn‟s post-hoc. For A and B, bold values indicate P<0.05. Note, no apparently significant (P<0.05) P-values were observed in post-hocs, with main effects non-significant.

| | Relative Abundance (mean ± SEM) | | | | | |
| --- | --- | --- | --- | --- | --- | --- |
| Genus (+/-Species) | Main Effect | Severe | Moderate | Mild | Asymptomatic | Exposed Control |
| *Alistipes* | 0.8119 | 0.0793±0.0166 | 0.0520±0.0084 | 0.0664±0.0196 | 0.0901±0.0272 | 0.0639±0.0095 |
| *Bacteroides* | **0.0075** | 0.2187±0.0272 | 0.2849±0.0305 | 0.0912±0.0226 | 0.2058±0.0606 | 0.1235±0.0178 |
| *Bifidobacterium* | **<0.0001** | 0.0018±0.0006 | 0.0037±0.0015 | 0.0507±0.0244 | 0.0840±0.0308 | 0.0755±0.0219 |
| *Blautia* | 0.2098 | 0.0495±0.0095 | 0.0426±0.0132 | 0.1037±0.0510 | 0.0260±0.0067 | 0.0261±0.0040 |
| *Clostridium* | 0.2721 | 0.0331±0.0062 | 0.0334±0.0055 | 0.0265±0.0083 | 0.0145±0.0025 | 0.0431±0.0075 |
| *Collinsella* | 0.7476 | 0.0145±0.0035 | 0.0107±0.0038 | 0.0203±0.0126 | 0.0334±0.0174 | 0.0146±0.0045 |
| *Dorea* | 0.4820 | 0.0218±0.0035 | 0.0126±0.0032 | 0.0174±0.0046 | 0.0152±0.0015 | 0.0137±0.0024 |
| *Eubacterium* | 0.9619 | 0.0436±0.0070 | 0.0367±0.0064 | 0.0366±0.0085 | 0.0332±0.0058 | 0.0441±0.0063 |
| *Faecalibacterium* | **0.0077** | 0.0209±0.0037 | 0.0428±0.0093 | 0.0359±0.0139 | 0.0597±0.0098 | 0.0550±0.0086 |
| *F. prausnitzii* | **0.0054** | 0.0220±<br>0.0041 | 0.0417±0.0092 | 0.0356±<br>0.0137 | 0.0589±0.0095 | 0.0542±<br>0.0085 |
| *Prevotella* | 0.1687 | 0.0149±<br>0.0118 | 0.0000±0.0000 | 0.0050±<br>0.0046 | 0.0023±0.0020 | 0.0110±<br>0.0086 |
| *Roseburium* | **0.0327** | 0.0146±<br>0.0037 | 0.0245±0.0075 | 0.0268±<br>0.0138 | 0.0274±0.0081 | 0.0329±<br>0.0056 |
| *Ruminococcus* | 0.8033 | 0.0384±<br>0.0076 | 0.0293±0.0073 | 0.0620±<br>0.0250 | 0.0394±0.0139 | 0.0376±<br>0.0079 |

**B.**
| Genus<br>(+/-Species) | P-value vs. exposed control |  |  |  | P-value vs. asymptomatic |  |  |
| --- | --- | --- | --- | --- | --- | --- | --- |
|  | Mild | Moderate | Severe | Asymptomatic | Mild | Moderate | Severe |
| *Alistipes* | >0.9999 | >0.9999 | >0.9999 | >0.9999 | >0.9999 | >0.9999 | >0.9999 |
| *Bacteroides* | >0.9999 | **0.0078** | >0.9999 | >0.9999 | >0.9999 | >0.9999 | >0.9999 |
| *Bifidobacterium* | >0.9999 | **0.0006** | **0.0026** | >0.9999 | >0.9999 | **0.0006** | **<0.0001** |
| *Blautia* | 0.4116 | >0.9999 | 0.6373 | >0.9999 | >0.9999 | >0.9999 | >0.9999 |
| *Clostridium* | >0.9999 | >0.9999 | >0.9999 | 0.4944 | >0.9999 | 0.9841 | >0.9999 |
| *Collinsella* | >0.9999 | >0.9999 | >0.9999 | >0.9999 | >0.9999 | >0.9999 | >0.9999 |
| *Dorea* | >0.9999 | >0.9999 | >0.9999 | >0.9999 | >0.9999 | >0.9999 | >0.9999 |
| *Eubacterium* | >0.9999 | >0.9999 | >0.9999 | >0.9999 | >0.9999 | >0.9999 | >0.9999 |
| *Faecalibacterium* | >0.9999 | >0.9999 | **0.0082** | >0.9999 | >0.9999 | >0.9999 | 0.1422 |
| *F. prausnitzii* | >0.9999 | >0.9999 | **0.0109** | >0.9999 | >0.9999 | >0.9999 | 0.1568 |
| *Prevotella* | >0.9999 | >0.9999 | >0.9999 | >0.9999 | >0.9999 | 0.8916 | >0.9999 |
| *Roseburium* | >0.9999 | >0.9999 | **0.0196** | >0.9999 | >0.9999 | >0.9999 | >0.9999 |
| *Ruminococcus* | >0.9999 | >0.9999 | >0.9999 | >0.9999 | >0.9999 | >0.9999 | >0.9999 |

Depicted in **Figure 3** are the 12 most abundant families and the 12 most abundant genera for patients, stratified by disease severity and in comparison to exposed controls. Distinguished by color, the bars represent the relative percent bacterial families and genera abundance. Note the reduced diversity of the microbiome of SARS-CoV-2 positive patients shown in column B.

**Figure 3:**
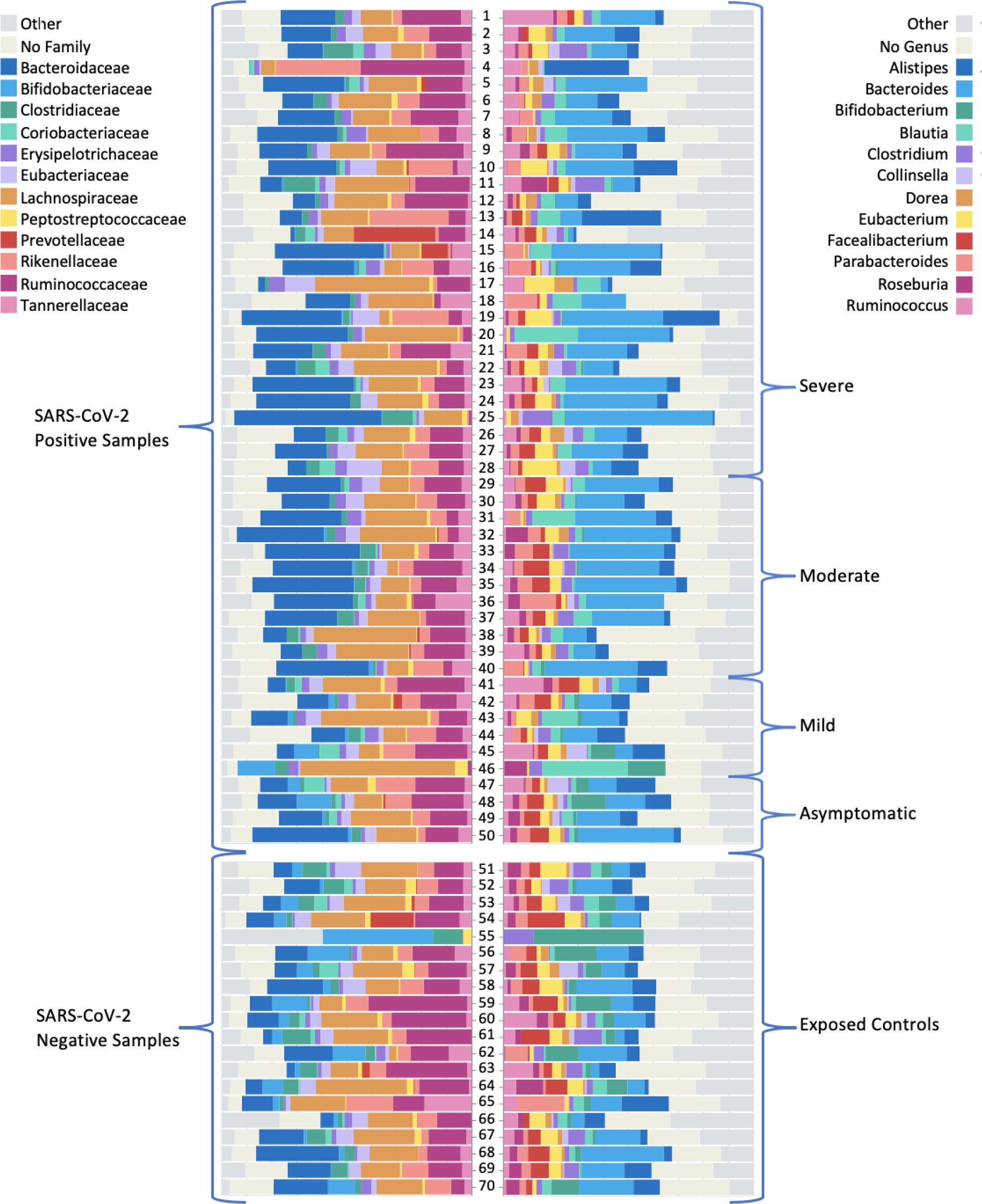
Graphic of relative abundance of the 12 most common A. families and B. genera. The top group represents the SARS-CoV-2 positive samples (n=50), stratified by severity. The bottom group represents the exposed control samples (n=20). The colored boxes represent the fraction of the entire rectangle composed of the given family/genera of bacteria.

**Figure 4** summarizes the microbiome changes according to SARS-CoV-2 positivity and severity, with green boxes depicting significant elevation and red boxes indicating significant depletion in genera / species abundance associated with SARS-CoV-2.

**Figure 4:**
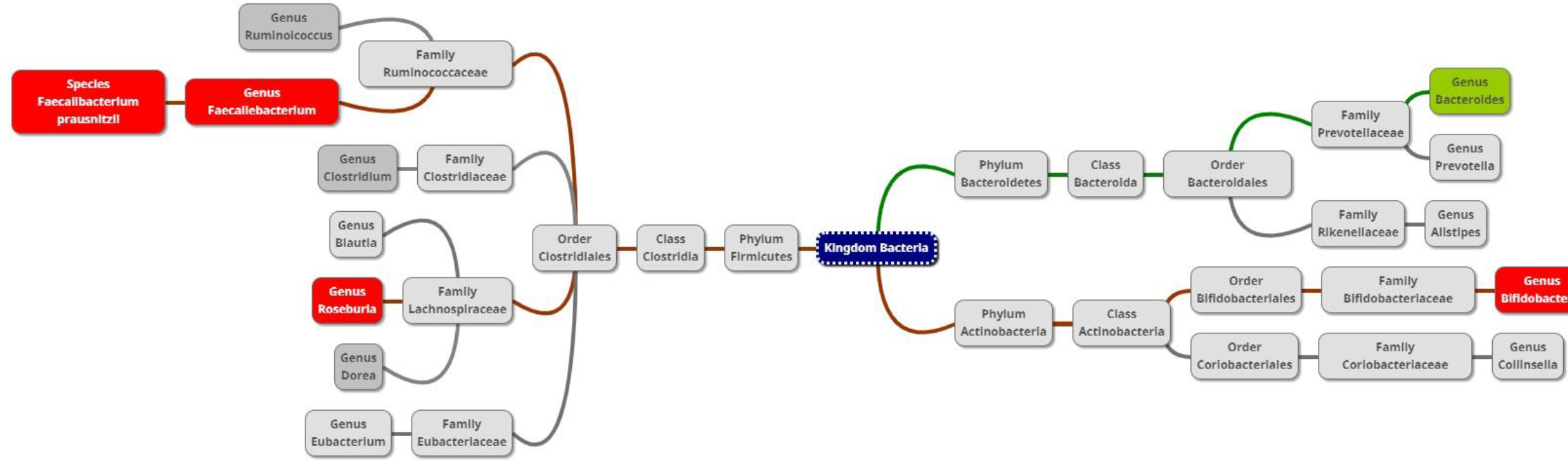
Diagram of taxa comparing the gut microbiome of SARS-CoV-2 patients and exposed controls. Red or green background indicates a significant depletion or increase (due to positivity or severity), respectively, of the genus or species in SARS-CoV-2 positive subjects.

**Figure 5A** and **5B** exhibit the relative abundance of *Bifidobacterium* for each subject, grouped by SARS-CoV-2 infection severity. This diagram, with subjects groups ordered by severity (from severe on left, to exposed controls on the right), depicts how Bifidobacterium abundance increases as severity decreases.

**Figure 5:**
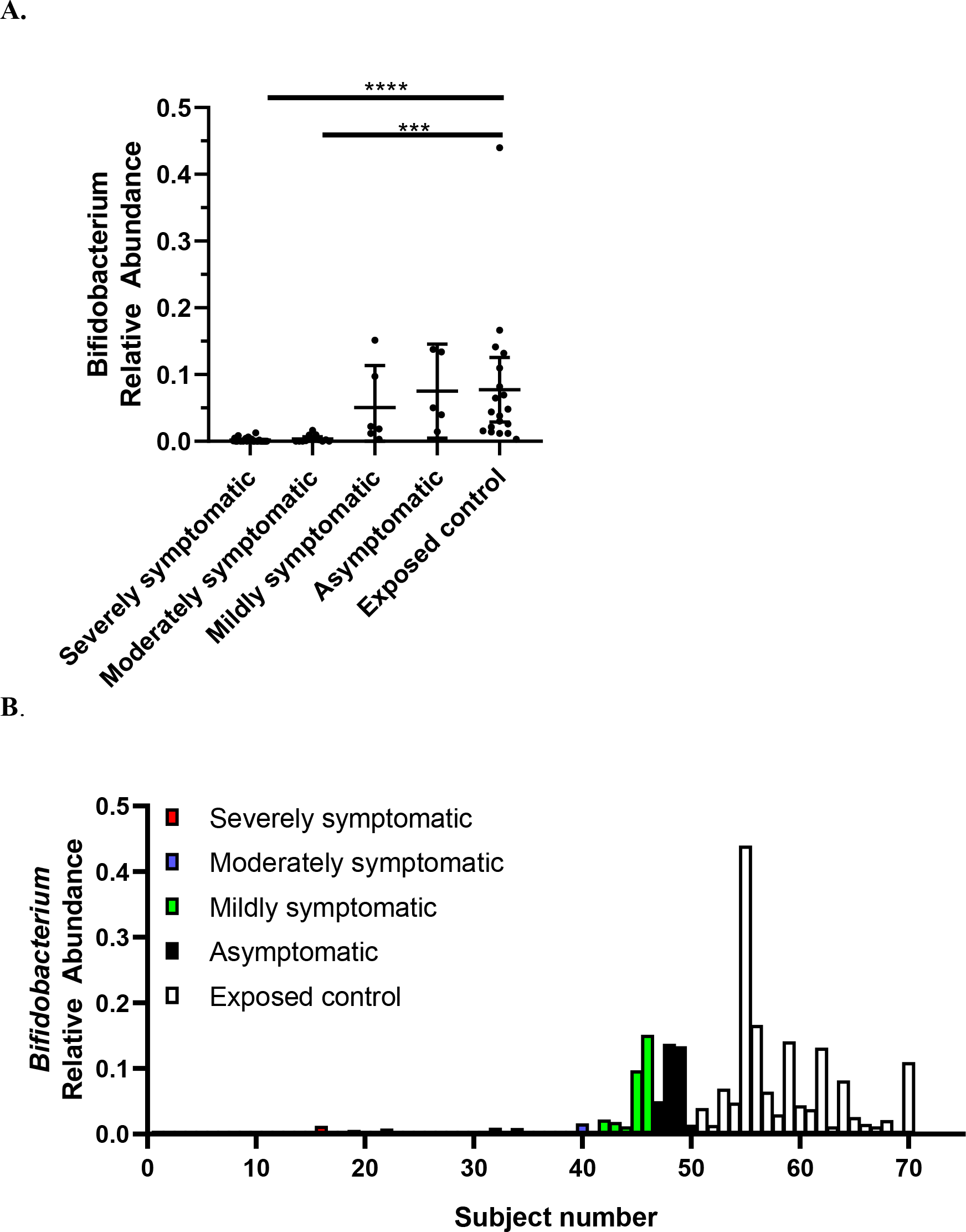
Relative abundance of *Bifidobacteria* in SARS-CoV-2 positive patients (n=50) vs. SARS-CoV-2 negative exposed controls (n=20). Data is plotted as A. mean with error bars for 95% CI and B. individual points of relative abundance for varying SARS-CoV-2 infection severity. Analyzed via Kruskal-Wallis test, there were significant reductions in *Bifidobacteria* relative abundance for severely (P<0.0001) and moderately (P=0.0002) symptomatic patients. Subjects 1-28 = severely symptomatic; subjects 29-40 = moderately symptomatic; subjects 41-46 = mildly symptomatic subjects; subjects 47-50 = asymptomatic; subjects 51 – 70 = exposed control. Note: Figure A and B depict same data.

Analysis of the beta-diversity of subjects demonstrated that the diversity of control subjects cluster separately from that of patients. **Figure 6A** shows the beta-diversity weighted (quantitative) UniFrac analysis featuring phyla bacterial profiles for all individuals in the study (n=70). **Figure 6A** reveals that, although there is a range of dissimilarity, the SARS-CoV-2 negative individuals are more similar to one another than they are to SARS-CoV-2 positive patients. The matrix also highlights clusters of similarity among SARS-CoV-2 positive patients, and darker quadrants of dissimilarity where positive and negative patients intersect. At a more granular level, **Figure 6B** utilized principal component (PC) analysis of genera where the axes depict the percent of variance. In PC analysis, points closer together are more similar (less divergence with axis representing directions of divergence). Herein the PC1 accounts for 43.16% of the variation, whereas PC2 accounts for 12.78%. This analysis reveals a clear divergence of a subset of SARS-CoV-2 positive patients clustering on the right side tracking along the x-axis (PC1), highlighting microbiota divergence as a function of disease. Thus, **Figure 6** shows that exposed controls cluster similar separately from SARS-CoV-2 patients; i.e. patients are more similar in terms of their microbiome to each other than to controls.

**Figure 6:**
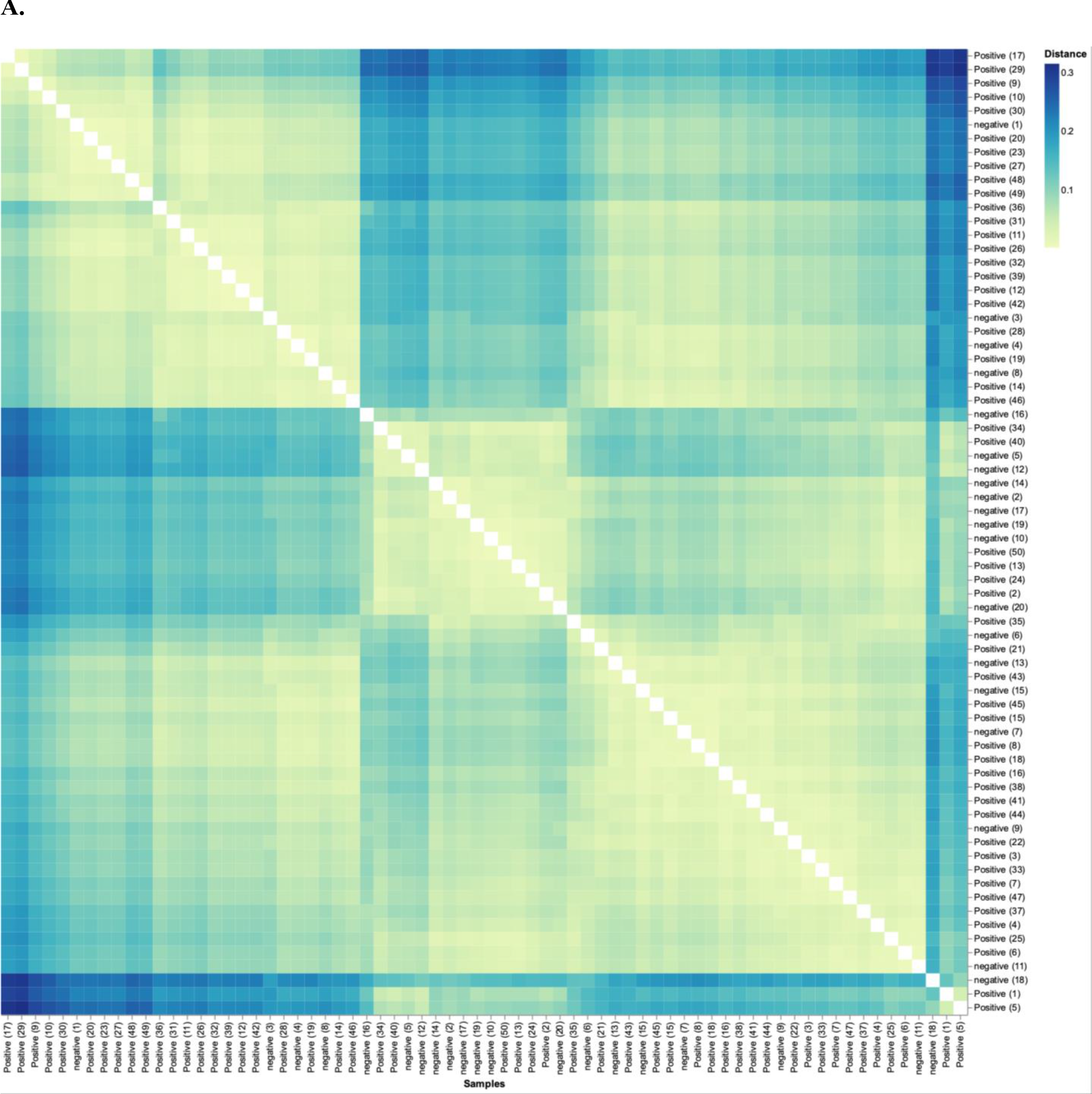
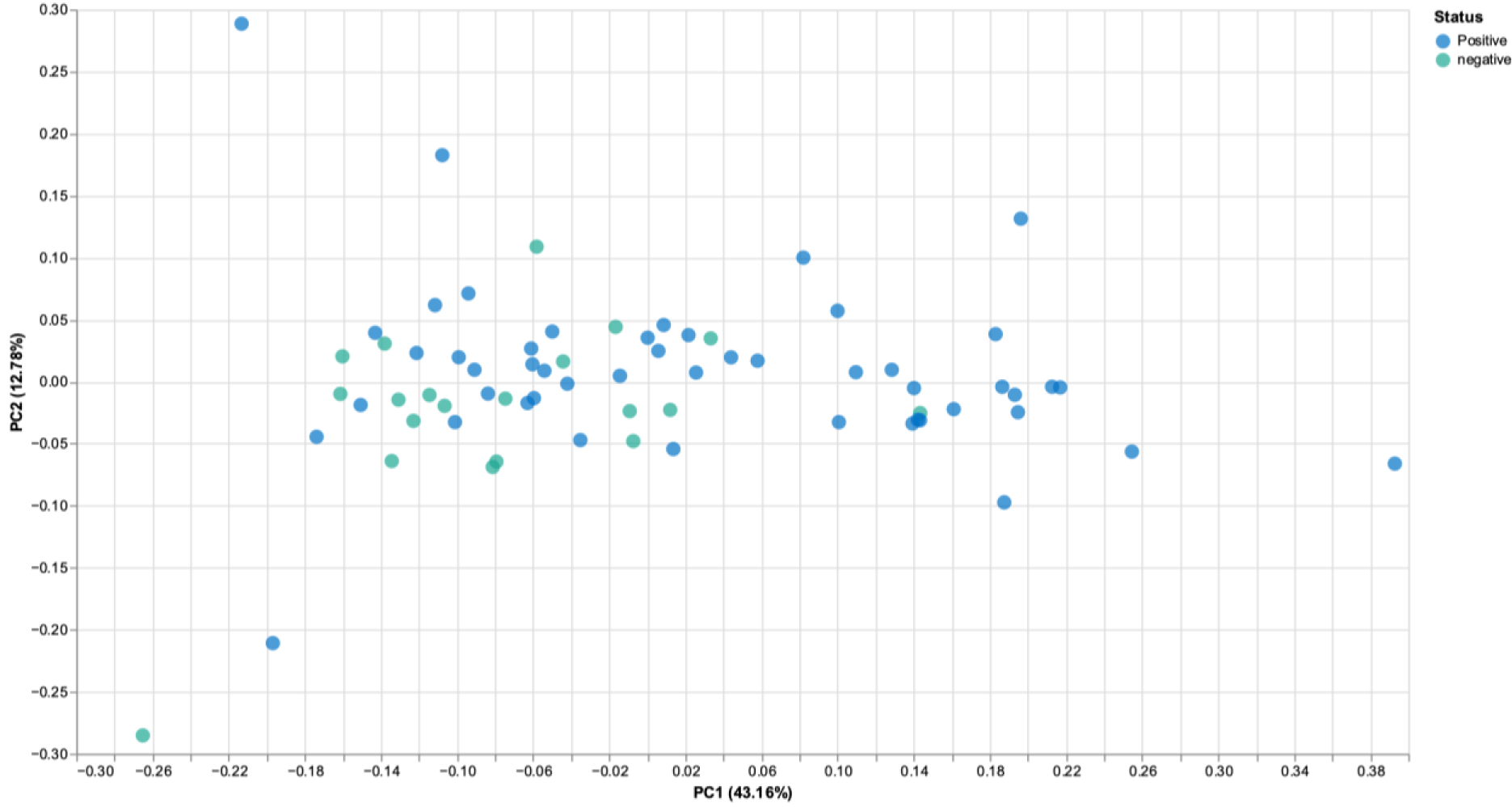
SARS-CoV-2 positive patients microbiome is more similar to each other than to that of exposed controls. A. Weighted UniFrac distance matrix of phylum level SARS-CoV-2 positive (n=50) and exposed negative control samples (n=20). Distance of microbiome differences increases with increasing blue color intensity (see legend top right). The center of the diagram consists of negative subjects on both axis, and is yellow indicative of less distance (i.e. less difference in microbiome). The central area of the left as well as central-top side of diagram, consists of negative subjects on one axis and positive on the other, and are darker blue, indicative of more distance (more difference in microbiome). B. Principal component analysis of microbiota from SARS-CoV- 2 positive (n=50) and exposed negative controls (n=20). Dots closer in distance are more similar in microbiome composition. Axes depict the percent of variance explained by principal component (PC) 1 and 2. Plots are based on bacterial genera relative abundance profiles.

## DISCUSSION

### Immune function and health could be enhanced by bacterial abundance

Interactions between the host and gut microbiota are complex, numerous, and bidirectional. Gut microbiota regulate the development and function of the innate and adaptive immune systems,[33] potentially allowing them to protect against infections and infection severity. The primary findings of our study are that SARS-CoV-2 positivity and infection severity are associated with decreased levels of the protective *Bifidobacterium* and *Faecalibacterium* genera and with decreased bacterial diversity, as exemplified by the Shannon and Simpson indices. This accords with studies showing bacterial diversity inversely relates to the presence of various common disorders.[34] Uniquely, our study compared SARS-CoV-2 exposed SARS-CoV-2 negative persons (i.e., controls) with symptomatic and asymptomatic SARS-CoV-2 positive patients. Thus, we controlled for SARS-CoV-2 exposure.

The genus *Bifidobacterium* has important immune functions,[8] is a major component of the microbiome and is frequently used in probiotics.[35] Bifidobacteria increase Treg responses and reduce cell damage by inhibiting TNF-α and macrophages.[36] Bifidobacteria also protect against intestinal epithelial cell damage independently from their effects on TNF-α production. The exopolysaccharide coat which is a feature of some *Bifidobacterium* plays a significant role in this protective effect.[37] These immune functions of *Bifidobacterium* could be critical in relation to its SARS-CoV-2 infection-prevention effects.

Evidence has accumulated to support a beneficial effect from supplementation with *Bifidobacterium* in numerous disease states.[38] The numbers of commensal Bifidobacteria have been shown to decrease with age and obesity, major SARS-CoV-2 infection risk factors. We demonstrate that patients with a more severe course of viral infection had decreased abundance of *Bifidobacterium*.

However, it should be noted that there are no definitive studies concerning what constitutes a normal baseline abundance of *Bifidobacterium* in a “healthy” individual.

The abundance of *Faecalibacterium* genus and *F. prausnitzii* species were also inversely related to SARS-CoV-2 positivity and SARS-CoV-2 infection severity in this analysis. Age and diabetes are risk factors for SARS-CoV-2 infection, and *F. prausnitzii* levels decline markedly in elder and diabetic populations.[38] In fact, *Faecalibacterium* levels have been considered an indirect “indicator” of overall human health.[39] The abundance of *F. prausnitzii* is reduced by the “Western” diet (consumption of more meat, animal fat, sugar, processed foods, and low fiber), while it is enhanced by the high-fiber containing “Mediterranean” diet of vegetables and fruits with low meat intake.[40] Preliminary studies showed that reduced ingestion of a Mediterranean diet within the same country is associated with increased SARS-CoV-2 related death rates.[41] In short, we show that *F. prausnitzii* levels negatively correlated to SARS-CoV-2 infection severity and prior studies show that reduced *F. prausnitzii* is associated with SARS-CoV-2 infection vulnerabilities such as age, diabetes, obesity, and possibly diet.

SARS-CoV-2 positivity and severity were also associated with decreased abundance of *Roseburium* and increased abundance of *Bacteroides.* The implications of these changes remain unclear.

### Innate immunity could be enhanced by increased level of beneficial bacteria

The pathological impact of SARS-CoV-2 infection includes both direct effects from viral invasion and complex immunological responses including, in its most severe form, the „cytokine storm.‟ The cytokine storm is the result of a sudden increase in circulating levels of pro-inflammatory cytokines produced by activated macrophages, mast cells, endothelial cells, and epithelial cells during innate immune responses, which appear to be modulated by the abundance of *Bifidobacteria* and *Faecalibacterium* and bacterial diversity(5, 23, 25) Steroid treatment has situational success in SARS-CoV-2 infection, based on suppressing this over activation of the innate immune system, reviewed by Tang et al.[42] Zhao et al. reported that elevated serum levels of pro-inflammatory cytokines such as IL-16 and IL- 17 predict poor prognoses in patients with SARS-CoV-2 infection.[43] Also, Tao et al. showed that changes in gut microbiota composition might contribute to SARS-CoV-2-induced production of inflammatory cytokines in the intestine, which may lead to cytokine storm onset.[29] Both authors report significantly reduced gut microbiota diversity and increased opportunistic pathogens in SARS-CoV-2 patients. Interestingly, the bloom of opportunistic pathogens positively correlated with the number of Th17 cells. Bozkurt et al. reported that IL-6 and IL-17 promote viral persistence by immune interactions through cellular autophagy via the inositol-requiring enzyme 1 pathway.[16] Additionally, some species of *Bifidobacterium* are likely to prevent the replication of coronaviruses by reducing endoplasmic reticulum stress, also through the inositol-requiring enzyme 1 pathway. Reduced *Bifidobacterium* abundance has been observed in the gut microbiome of patients with IBD, which has mechanisms involving IL-17.[31] Furthermore, the direct endoscopic delivery of *Bifidobacterium* has been shown to be effective in promoting symptom resolution and mucosal healing in IBD―an effect likely to be associated with the anti-Th17 effect of *Bifidobacterium.*(8) **Figure 7** demonstrates how *Bifidobacteria* might hypothetically quell a heightened immune response by dampening the effect of the master switch TNF-α.

**Figure 7:**
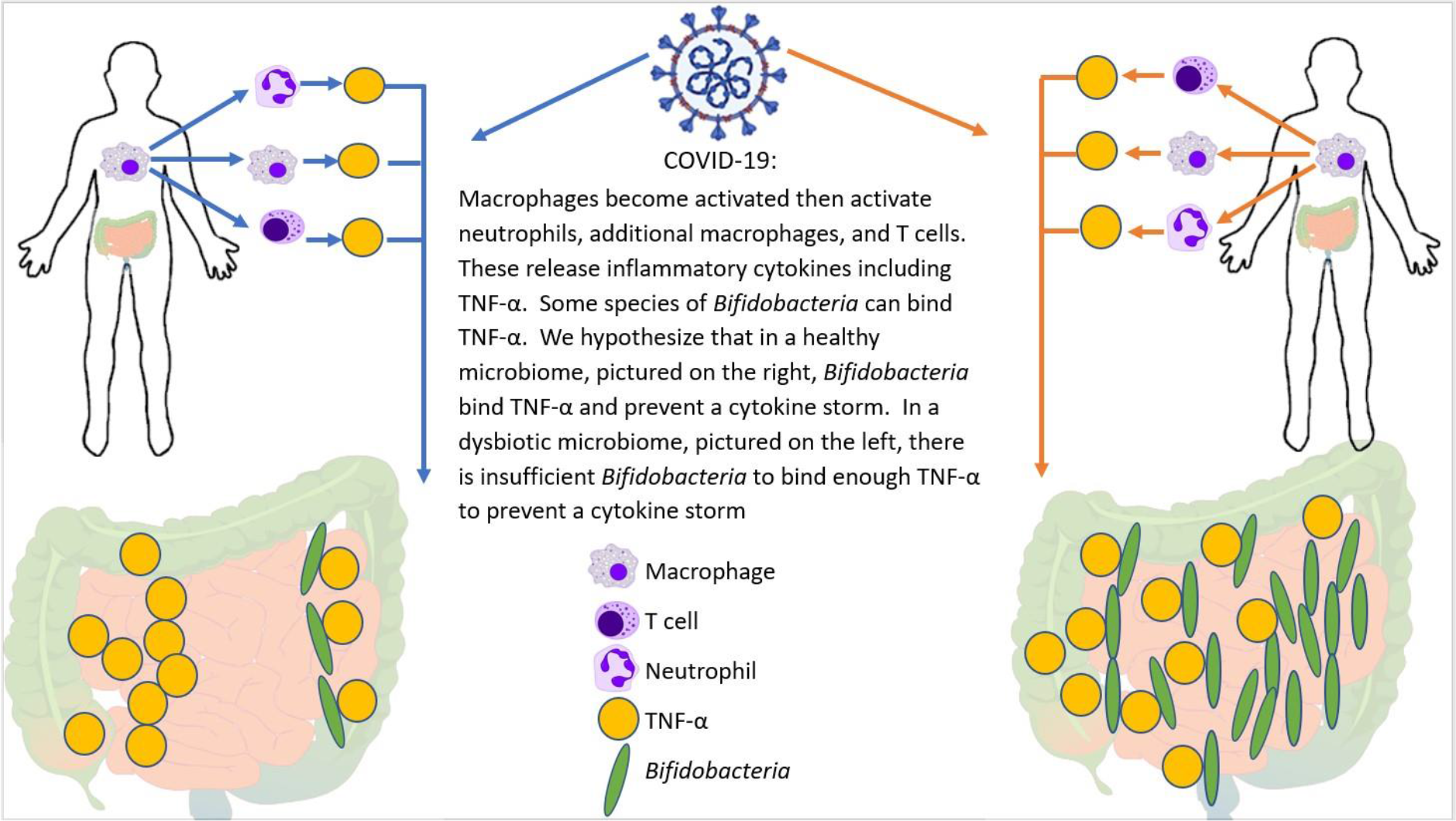
Potential mechanism for cytokine storm and immune hyper-response in SARS-CoV-2 positive patients. In individuals infected with SARS-CoV-2, the macrophages become activated; these in turn activate T-cells, additional macrophages, and neutrophils―all of which release cytokines, including TNF-α. *Bifidobacteria*, when present in sufficient numbers, can bind to TNF-α and prevent the subsequent cytokine storm. Therefore, patients with a bifidobacterial dysbiosis characterized by low levels of *Bifidobacteria* lack this line of defence, which may lead to a cytokine storm.

## CONCLUSIONS

Given our cross-sectional study design, it is not possible to determine whether the differences in *Bifidobacterium* levels observed between patients and exposed controls preceded or followed infection. If preceding infection, they could be a marker of susceptibility and boosting *Bifidobacterium* levels might decrease the risk or severity of SARS-CoV-2 infection. If these changes followed infection, alteration of the gut microbiome (such as through fecal microbiota transplantation or possibly probiotic supplementation) to increase *Bifidobacterium spp.* could be an area worth exploring for improved outcomes. If future studies can demonstrate improved outcomes, such therapy can be considered for complex cases of SARS-CoV-2 infection, such as “long-haulers,” and those with severe disease. Developing outbreaks within tightly closed communities such as nursing homes might be a good setting in which to assess susceptibility: fecal samples could be collected during the outbreak and run *post hoc* on “cases” and “controls.” Future studies of individuals with baseline pre-pandemic microbiome data would be highly valuable, although acquiring such baseline pre-infection microbiome data is still costly.

With the lack of data on the gut microbiome prior to onset of SARS-CoV-2 infection, we cannot completely rule out the confounding effect of illness on the microbiome. Nonetheless, we eliminated effects of treatment on the gut microbiome by sampling prior to administration of SARS-CoV-2 infection therapeutics of any kind and within 48 hrs. of symptom onset. Specifically, no subjects were given antibiotics, antivirals, anti-inflammatory medicines, oxygen, or any other therapeutic agent between symptom onset or PCR positivity, and stool sampling. We also note that the prevalence of appetite changes, alterations of stool frequency, and GI symptoms, in general, were not significantly different between any of the severity groups or controls (Table 1), although the small sample sizes for some groups should be considered in evaluating these statistics.

SARS-CoV-2 infection presentation variability correlates with colon microbiome bacterial composition and overall diversity. The same changes we observe due to SARS-CoV-2 infection, namely reduced *Bifidobacterium* and/or *Faecalibacterium* abundance, are associated with SARS-CoV-2 infection risk factors including old age, obesity, and diabetes.[9, 38, 40, 44] Thus, colon microbiome diversity and relative abundance of *Bifidobacterium and Faecalibacterium* should be explored as potential markers for predicting SARS-CoV-2 infection severity.

In summary, we demonstrate in a study of PCR-positive and PCR-negative SARS-CoV-2-exposed subjects, reduced bacterial diversity and reduced levels of various genus / species are highly associated with both SARS-CoV-2 positivity and SARS-CoV-2 infection severity. These findings suggest that probiotic supplementation or fecal microbiota transplantation should be explored as a potential therapeutic avenue, for SARS-CoV-2 patients. Additionally, individual colon microbiome evaluation may predict vulnerability to the development of severe SARS-CoV-2 infection. Lastly, our data suggest a new area for exploration: if SARS-CoV-2 severity is found to be dependent on the microbiome, then accounting for microbiome differences could reduce variability in outcomes for SARS-CoV-2.

## ABBREVIATIONS

CDC (Center for Disease Control); GCP (good clinical practice); IBD (inflammatory bowel disease); ICH (International Council for Harmonisation); IL (interleukin); IRB (Independent Review Board); NGS (next generation sequencing); NIH (National Institute of Health); PC (Principal Component); PCR (polymerase chain reaction); PPE (personal protective equipment); SARS-CoV-2 (Coronavirus, COVID, COVID-19); TNF (tumour necrosis factor)

## DECLARATIONS

### Ethics approval and consent to participate

The study was conducted in accordance with ethical principles of the Declaration of Helsinki, the International Council for Harmonisation (ICH) Harmonised Tripartite Guideline for Good Clinical Practice (GCP), and the Ethical and Independent Review Board (IRB, #IRB00007807). All patients provided written informed consent to participate.

### Availability of data and materials

The datasets used and analyzed during the current study are available from the corresponding author on reasonable request.

### Authors’ contributions

All authors (SH, NS, HB, SD, AJP, JD, BDB, EMMQ, and TJB) participated in the drafting, critical revision, and approval of the final version of the manuscript. SH led study design. SH and AJP conducted the bioinformatic analysis. SD conducted the statistical analysis and was a major contributor to writing the paper. SH was primarily responsible for interpretation of the study results, with contributions from all authors. EMMQ and TJB are senior authors who provided overall direction and advice.

## Data Availability

Data available upon reasonable request from corresponding author, Dr. Sabine Hazan.

## Acknowledgements

Medical writing assistance was provided by Sonya Dave, PhD (an author on the publication) and was funded by ProgenaBiome. The authors thank all clinicians for their involvement and contribution to the study. The authors thank Kate Hendricks, MD, MPH&TM for many helpful editorial suggestions. Finally, the authors owe a depth of gratitude to the late Sydney M Finegold, MD for mentorship that sparked the interest in the microbiome to many scientists, including authors of this paper.

---

**Table 1.**
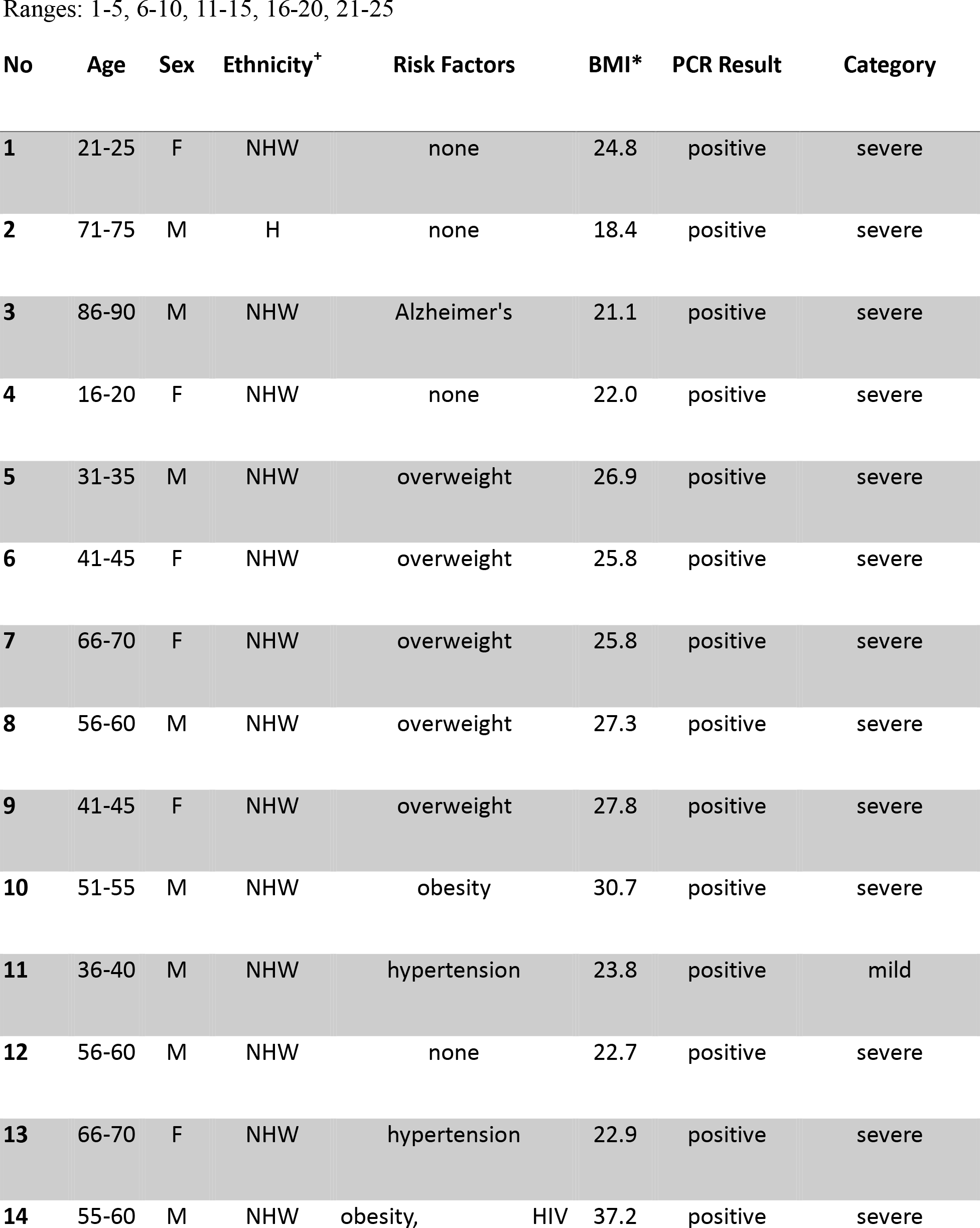
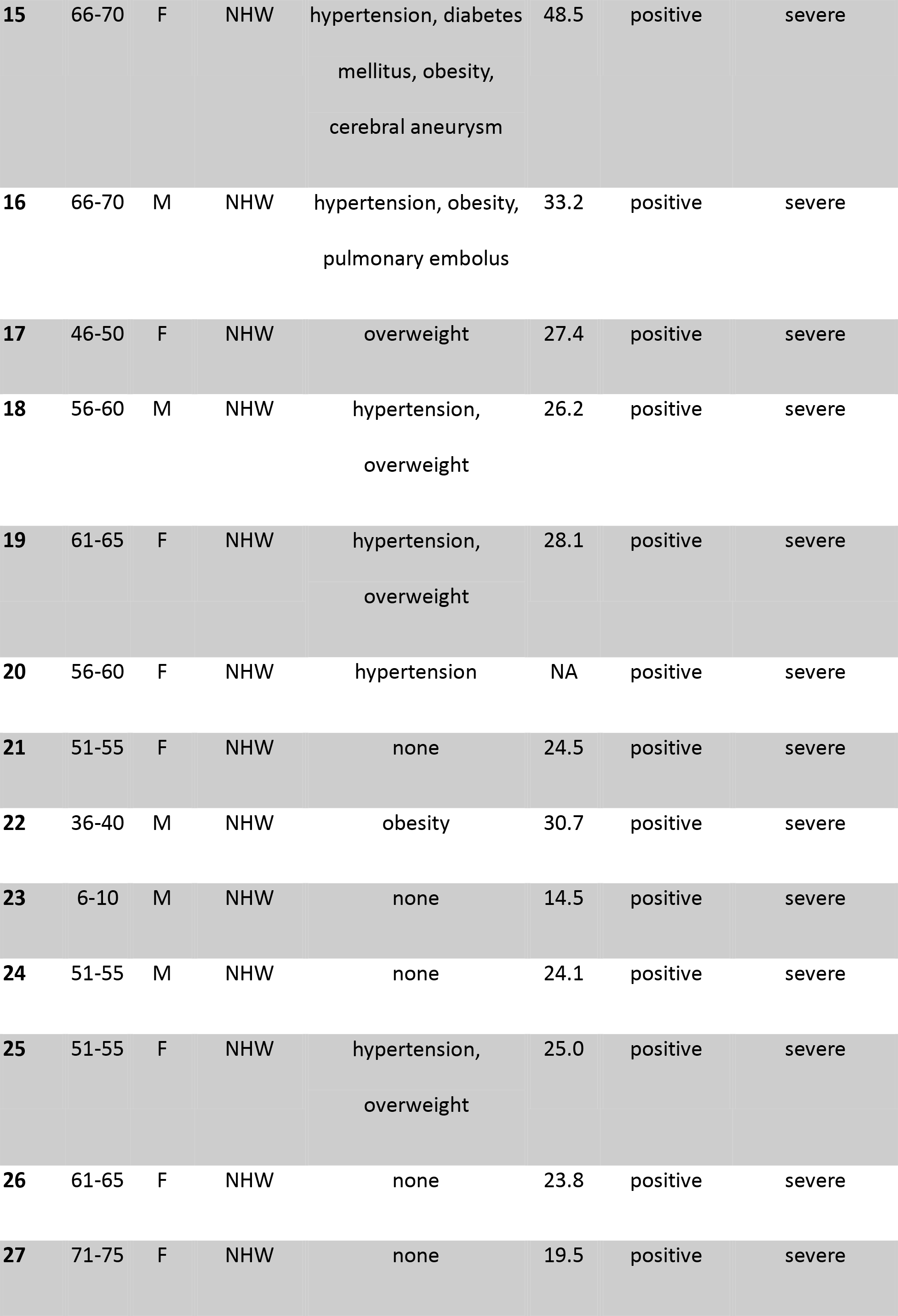
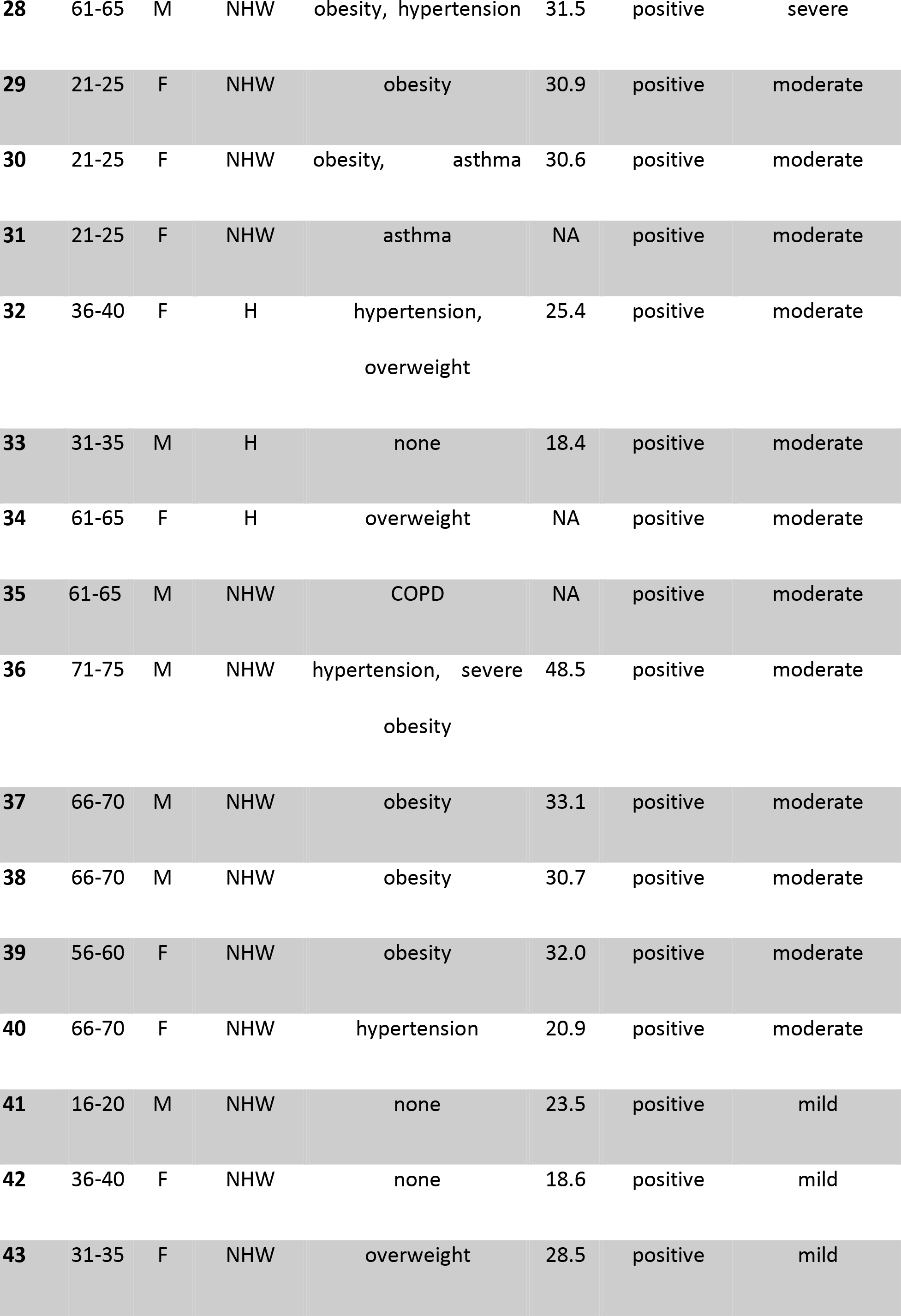
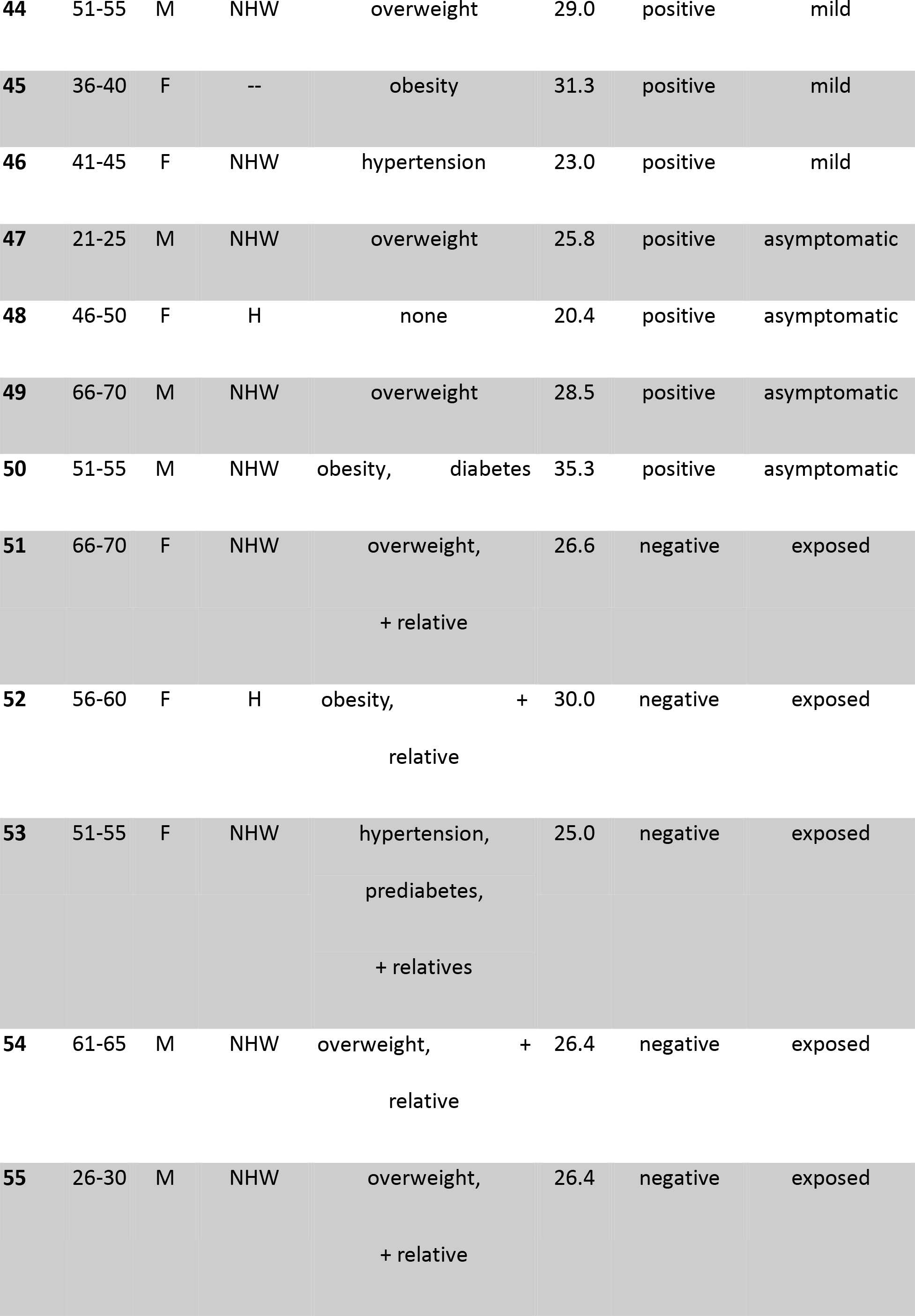
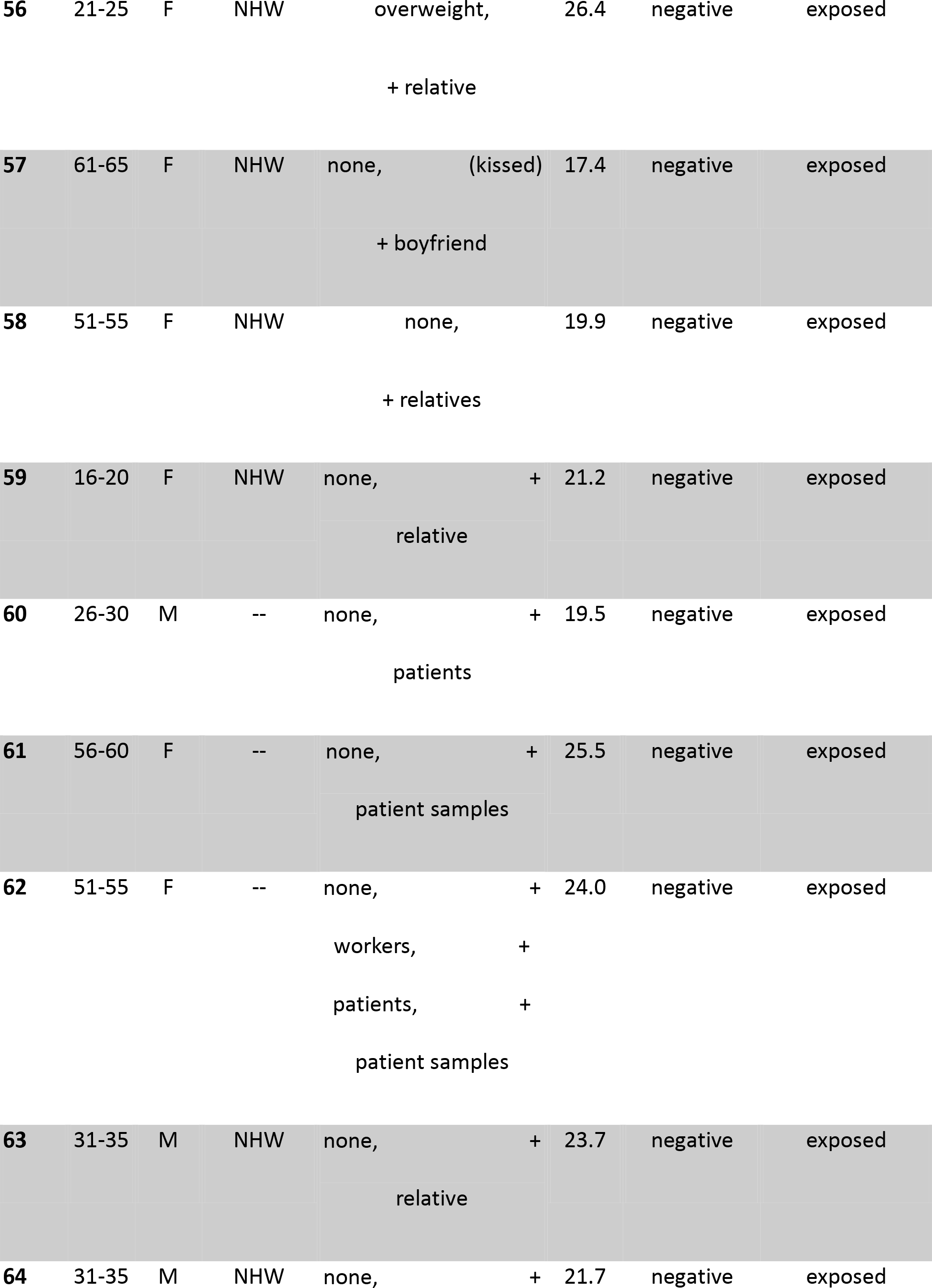
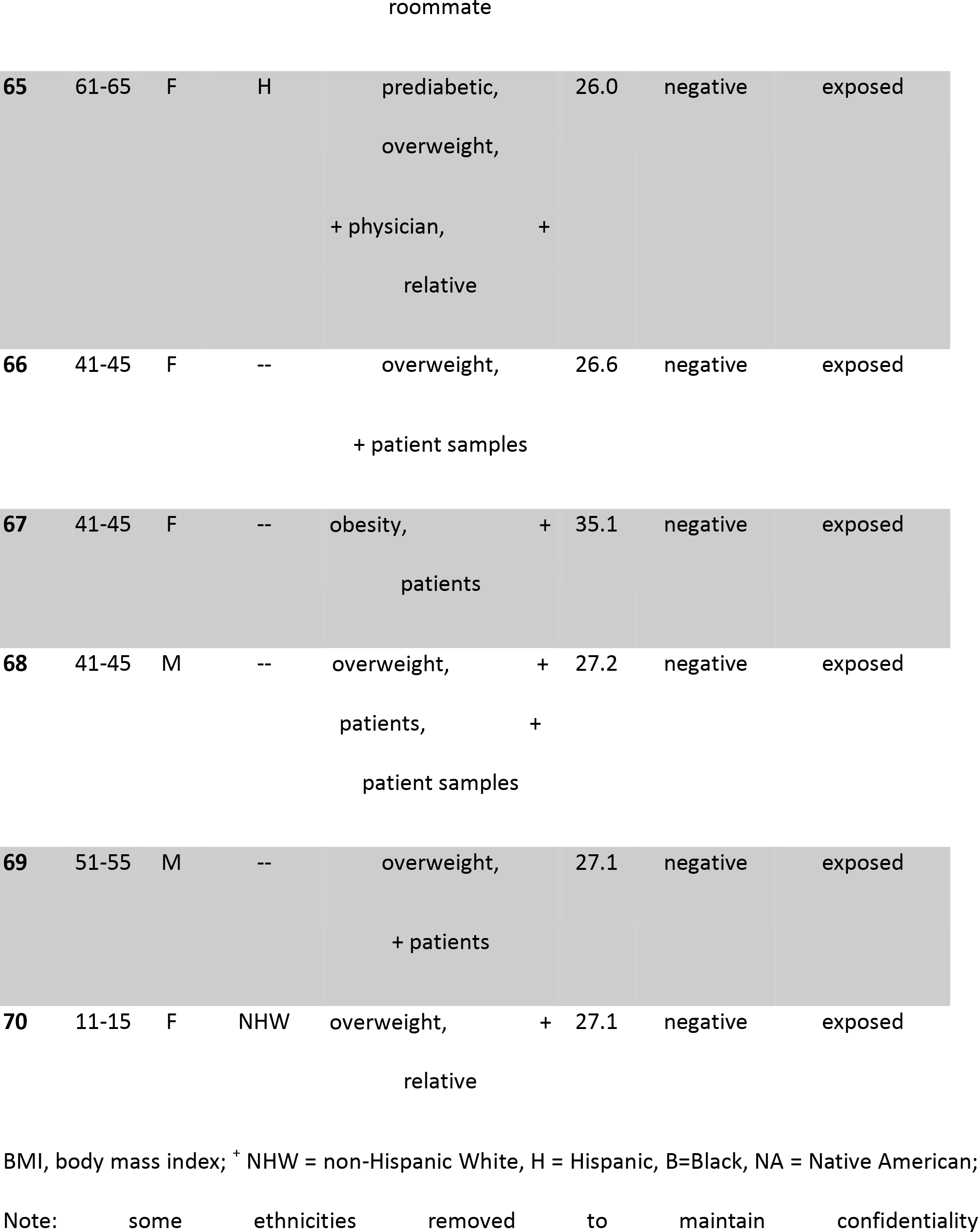
Demographic and baseline clinical characteristics of SARS-CoV-2-positive patients and SARS-CoV-2-negative exposed controls

**Supplementary Table S2:**
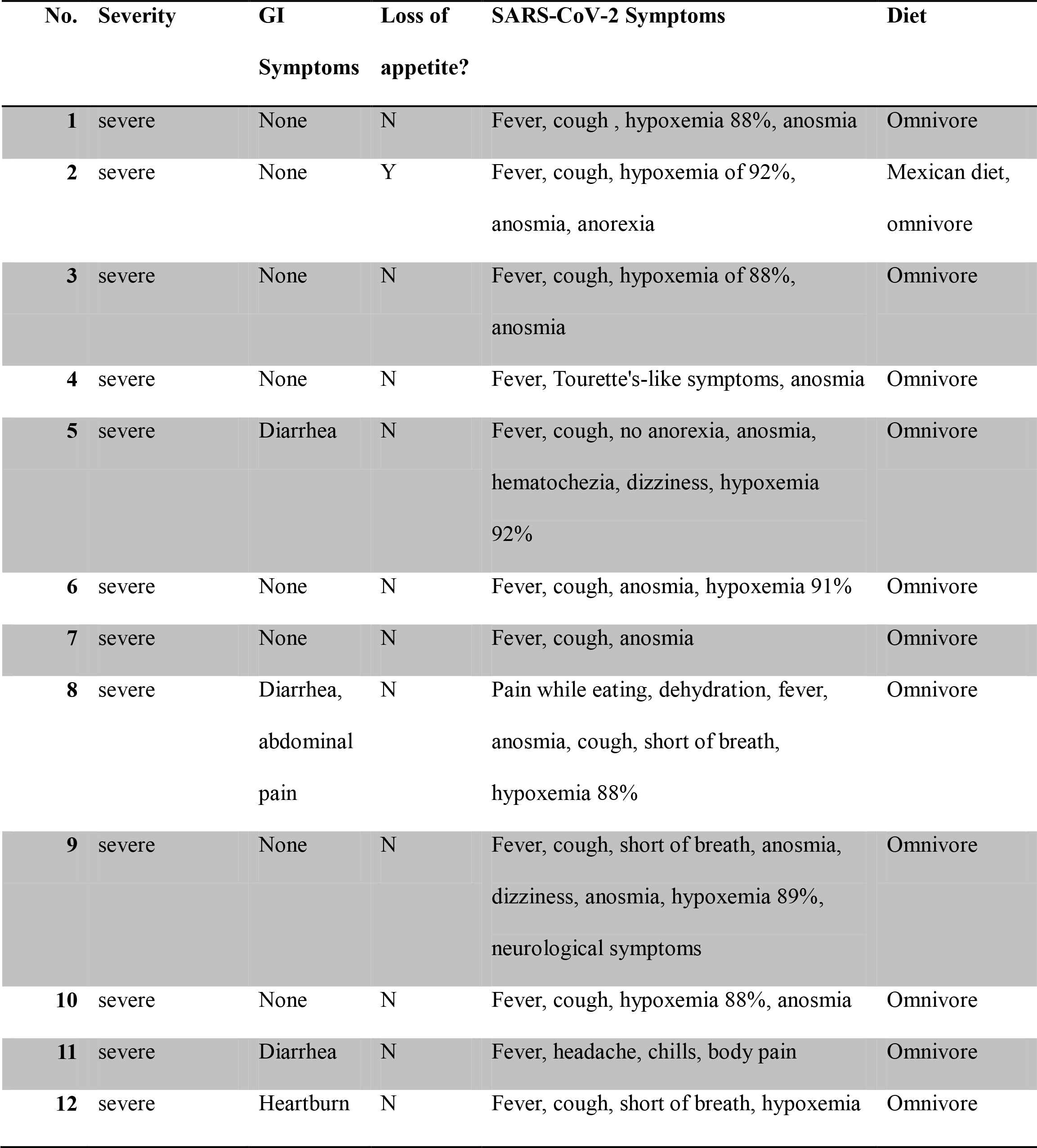
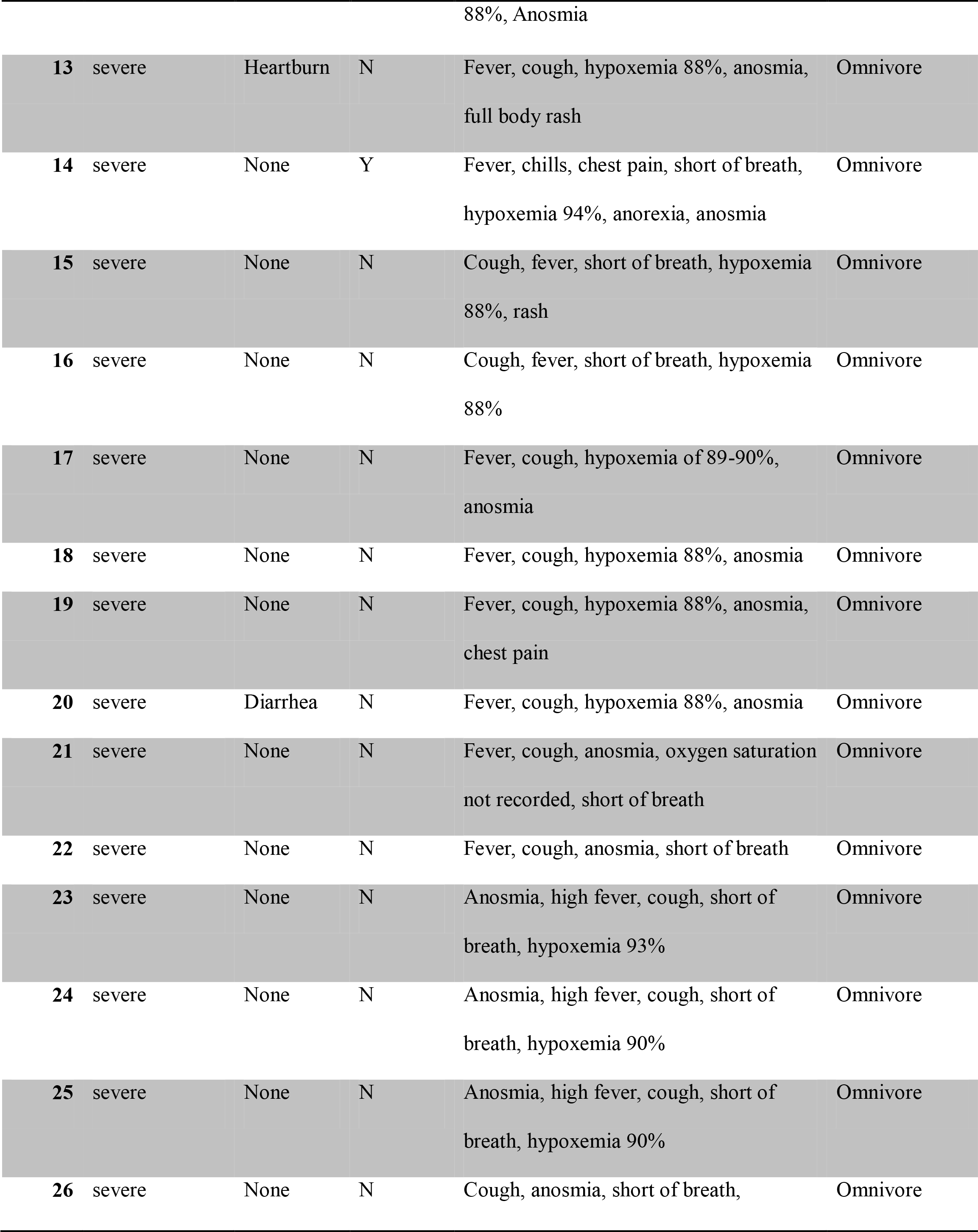
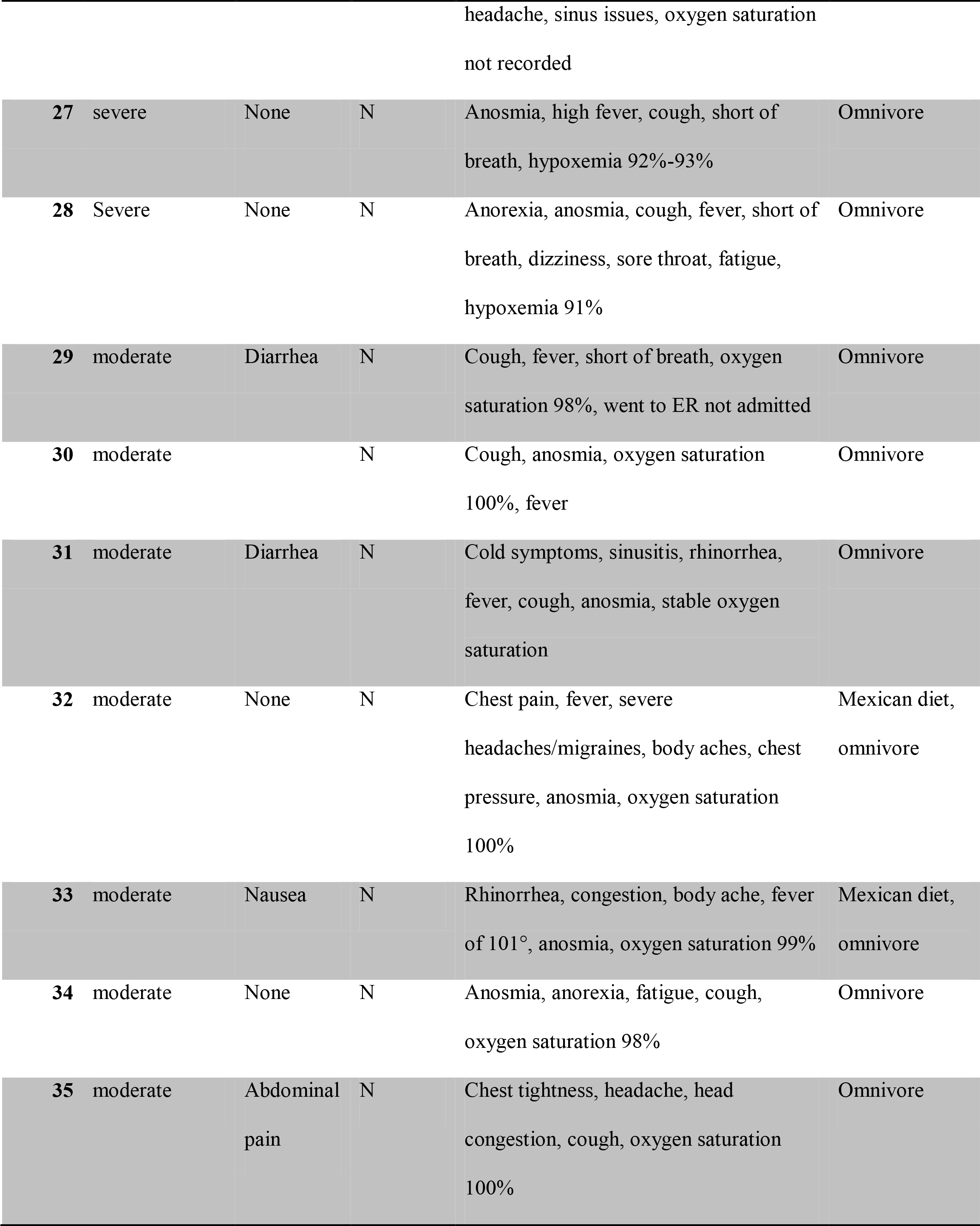
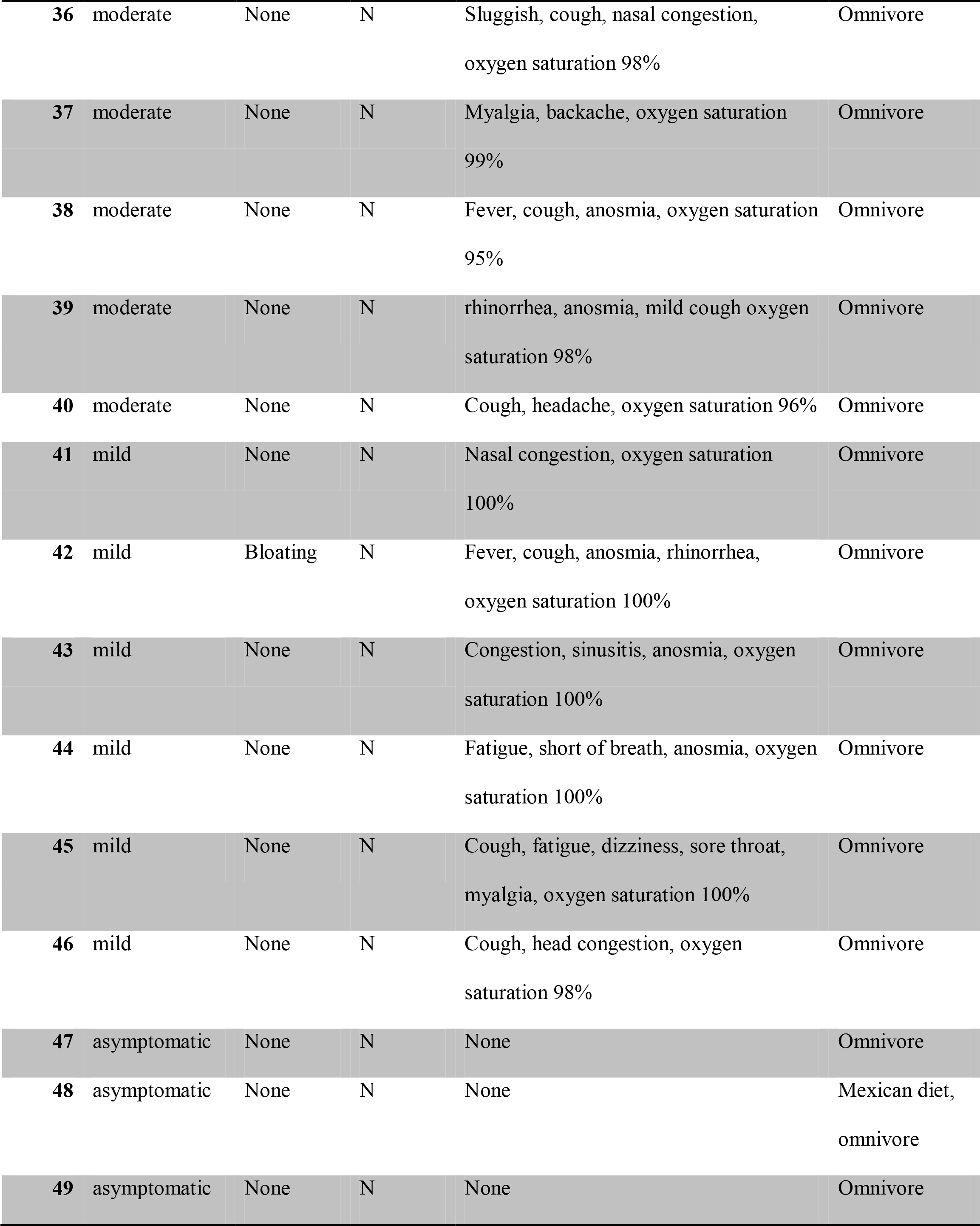
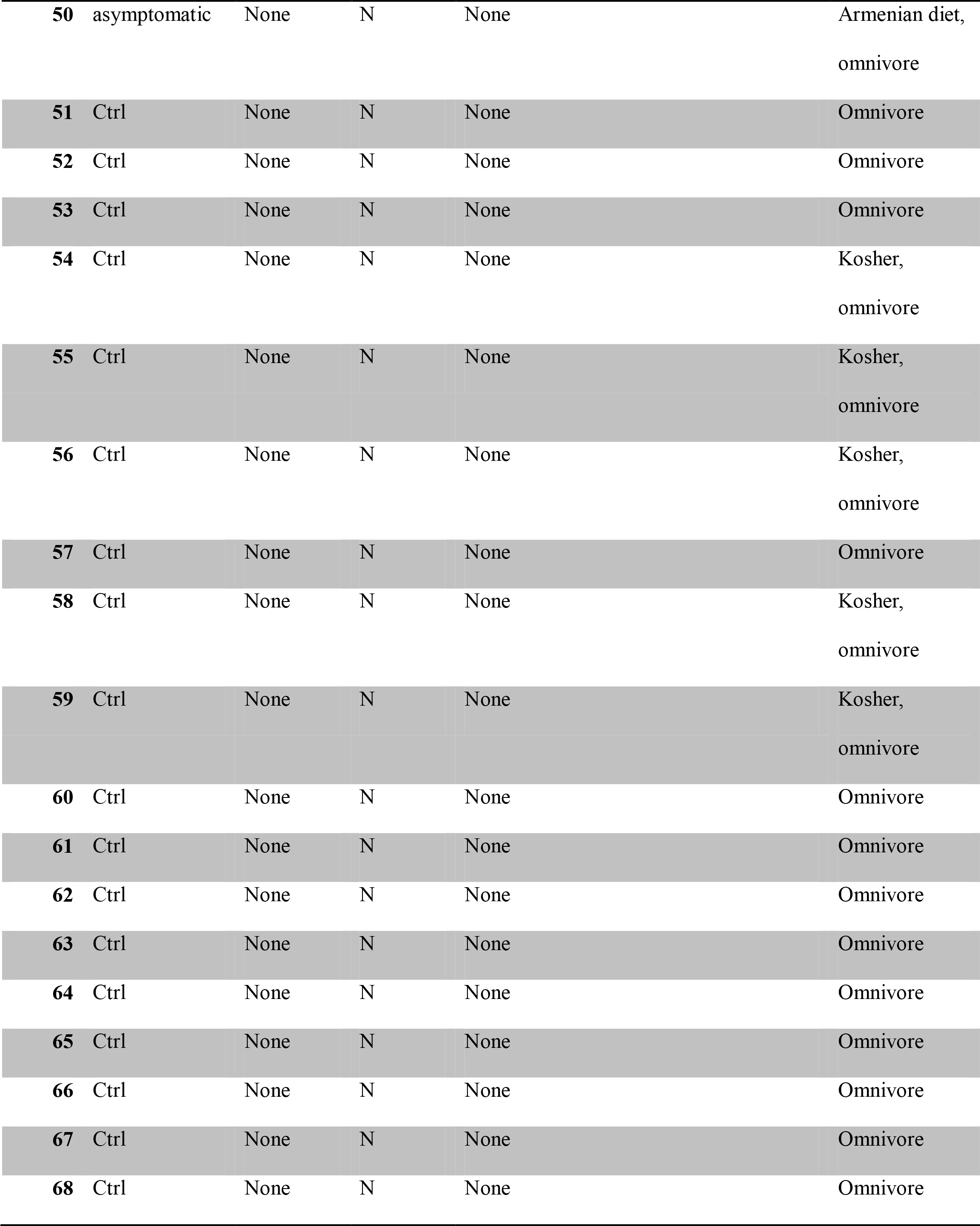
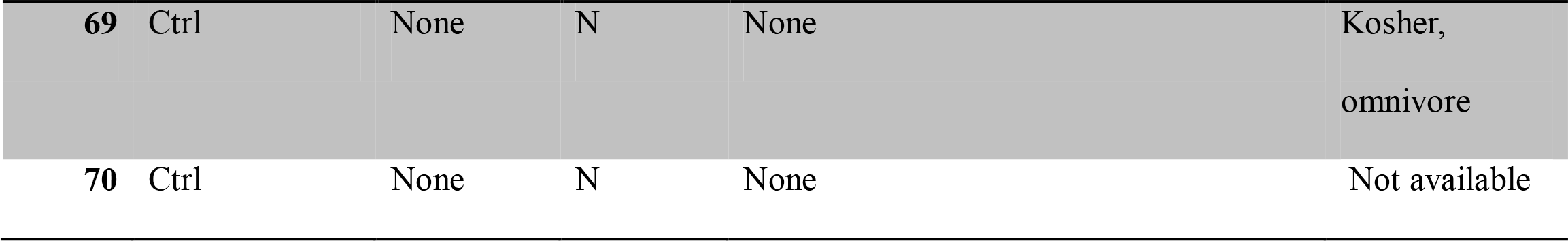
Detailed SARS-CoV-2 clinical characteristics and typical dietary habits of subjects.

